# Psychosocial Health Inequalities and Socioeconomic Deprivation Gradients Among Preschool Children in Care and Not in Care: An Administrative Health Data Study

**DOI:** 10.64898/2026.08.25.26361327

**Authors:** Daniel R. R. Bradford, Yara Abou Saab, Alex D. McMahon, Alastair H. Leyland, Mirjam Allik, Denise Brown

**Affiliations:** MRC/CSO Social and Public Health Sciences Unit, University of Glasgow; School of Health and Wellbeing, University of Glasgow; Dental School, University of Glasgow

## Abstract

**Importance:** Peschool children in care are at high risk for psychosocial health concerns. Population-based evidence is limited.

**Objective:** Estimate prevalence of psychosocial health concerns in children in care and not in care, and assess care-status differences stratified by deprivation.

**Design:** Population-based cross-sectional study using linked, routinely collected 27–30 Month Health Review and Scottish Birth Record data from April 2013 to March 2023.

**Setting:** Universal health review program in Scotland.

**Participants:** 7887 children in care and 445 547 children not in care.

**Exposures:** Care status at review, classified as in care or not.

**Main Outcomes and Measures:** Four outcome categories (emotional, behavioral, and/or attentional; personal and/or social; speech, language, and/or communication; and other developmental concerns) plus an aggregate indicator of any of the four. We estimated adjusted odds ratios comparing children in care with children not in care and examined whether these varied across deprivation groups. Models included deprivation group and adjusted for sex, age, and ethnicity.

**Results:** Psychosocial health concerns were more common in children in care (2290 of 7887; 29.0%) than children not in care (77 836 of 445 547; 17.5%; relative risk 1.66). Concerns were more common in children in care across all outcomes; the adjusted odds ratio for any recorded concern was 1.86 (95% CI, 1.77–1.96). Adjusted odds ratios varied by outcome from 1.57 (95% CI, 1.49–1.66) for speech, language, and/or communication concerns to 2.49 (95% CI, 2.34–2.66) for emotional, behavioral, and/or attentional concerns. Relative inequities between children in care and not in care decreased with increasing deprivation: adjusted odds ratios were 1.58 (95% CI, 1.45–1.72) in the most deprived fifth of areas and higher at 2.61 (95% CI, 2.25–3.03) in the least deprived fifth of areas. Prevalence of any recorded concern increased with deprivation in both care groups. The relative risk comparing the most deprived with the least deprived fifths of areas was 1.46 (95% CI, 1.29–1.66) among children in care and higher at 2.34 (95% CI, 2.29–2.40) among children not in care.

**Conclusions and Relevance:** Psychosocial health inequities are evident at age two between children in care and not in care, and vary with deprivation. Support for children in care and children living in more deprived areas should be prioritized.

## 1 Introduction

Children in the formal care of the state tend to have poorer psychosocial health than children not in care. This includes difficulties in domains such as language, behavior, and interpersonal skills [1–5]. Longitudinal evidence suggests differences persist, and may widen, across childhood [6]. Yet, evidence about this group is limited at the population level. We estimated psychosocial health concern prevalence at age two for children in care and not in care using routinely collected health data, and assessed whether the care-status differences varied with area-level socioeconomic deprivation.

Although being in care often represents a positive intervention, it is typically a signal of accumulated adversity. Two adverse factors are particularly relevant to the health of young children in care: abuse and neglect, and socioeconomic deprivation. Abuse and neglect are common reasons for entering care [2, 6–12]. Early maltreatment can disrupt attachment and psychological regulation, increasing emotional, behavioral, and developmental concerns, and can affect neurocognitive development [13, 14]. Children in care are also more likely to live in deprived areas [15–20], with increasing deprivation associated with poorer psychosocial outcomes in young children regardless of care status [21, 22]. Care experience and exposure to socioeconomic disadvantage co-occur and jointly shape health, but whether the association between care status and psychosocial health varies with deprivation is currently unclear.

Existing research suggests a substantial proportion of preschool children in care have psychosocial health issues across multiple domains [2, 23–26]. Speech, language, and/or communication (SLC) difficulties also appear more common among children in care across age groups [3, 27]. However, comparator groups are often absent from studies of psychosocial health in young children in care [28], limiting assessment of the extent to which concern prevalence differs from that among children not in care. Some studies have also included disproportionate numbers of children living in areas of high deprivation, potentially conflating patterns associated with care status and deprivation [29]. Some contrasting findings exist, including lower SLC concern prevalence among children in out-of-home care in a Finnish birth cohort spanning all ages [30]. Population-level prevalence estimates, together with relative and absolute differences, can help clarify whether specific aspects of psychosocial health require attention in particular population subgroups.

Because care status and area deprivation capture related but distinct forms of adversity, the association between care status and psychosocial health may differ across deprivation groups. Examining deprivation gradients separately by care status shows whether socioeconomic inequalities differ between children in care and children not in care. Together, these different but complementary comparisons are important for beginning to characterize how care status and deprivation are associated with psychosocial health. Examining these differences at age two is also important to show show whether inequalities are already apparent in early childhood. To our knowledge, no studies have quantitatively examined the interaction of care status and deprivation. We addressed this using population-level health data from the universal NHS Scotland 27–30 Month Health Review linked to area-level deprivation data [31, 32]. Previous studies have used this review to study early development with some attention to deprivation-related differences, but not differences between children in care and children not in care [33, 34].

### 1.1 Study Aims

We aimed to quantify the prevalence of psychosocial health concerns among children in care and children not in care using population-wide data (RQ1). We also aimed to examine whether differences in psychosocial health varied by area-level socioeconomic deprivation. We examined the interaction of care status and deprivation from two complementary perspectives. First, we compared children in care with children not in care within each deprivation group (RQ2). Second, we examined the deprivation gradient separately among children in care and children not in care (RQ3). The research questions were:

1. Does psychosocial health differ between two-year-old children in care and children not in care?
2. Does the association between care status and psychosocial health vary with area-level socioeconomic deprivation in two-year-old children?
3. Does the socioeconomic deprivation gradient in psychosocial health differ between two-year-old children in care and children not in care?

## 2 Methods

### 2.1 Participants

We included children in Scotland who underwent a 27–30 Month Health Review (described below) on or between April 1, 2013, and March 31, 2023 and were aged two years (104–156 weeks). This wider age range allowed for routine and expected variation in review timing, including reviews conducted outside the 27–30-month window because of parental and health visitor availability, while retaining reviews with plausible age data. Each child contributed one review. We included all eligible records in the available extract; no sample-size calculation was performed.

### 2.2 Data Sources

We obtained health and demographic data from completed 27–30 Month Health Reviews conducted by NHS Scotland as part of the Child Health Systems Programme [35]. Clinicians carry out these reviews as holistic assessments of a child’s health and development. Review coverage is high and varies only modestly by deprivation group, from 88.7% in the most deprived fifth of areas to 89.8% in the least deprived fifth during the study period. Review data also include Read v2 clinical codes, a coded clinical terminology used in UK primary and community health records. We used Scottish Index of Multiple Deprivation (SIMD) data to measure area-level socioeconomic deprivation. SIMD ranks small geographic areas, known as data zones, from most to least deprived. Although SIMD includes health-related measures, prior work suggests this does not affect substantive conclusions when estimating socioeconomic health inequalities [36, 37]. We used Scottish Birth Record data to improve ethnicity completeness. Health review data were assigned the temporally nearest SIMD version based on the child’s place of residence at review, with 2013 reviews assigned to SIMD2012, 2014–17 reviews to SIMD2016, and 2018–23 reviews to SIMD2020v2 [38]. Public Health Scotland’s Electronic Data Research and Innovation Service (eDRIS) carried out all linkage and provided the study team access only to a pseudonymized linked dataset. Details of linkage methods and linkage-quality metrics were not provided to the study team. Access to the health data was granted by the Public Benefit and Privacy Panel for Health and Social Care (reference 2021-0199). Ethics approval and informed consent were not required for this secondary analysis of routinely collected health data.

### 2.3 Measures

#### 2.3.1 Exposure

The exposure was care status at the time of the health review. Children in care included those recorded as in state care while looked after at home, in kinship care, foster care, living with prospective adopters, other community placements, or residential care. We derived care status primarily from the structured care-status field and supplemented this with Read v2 codes indicating that a child was in care (see Table A6; N = 42). We classified care status as in care, not in care, or unknown. The latter group was retained in all analyses but our results focus on children in care versus children not in care.

#### 2.3.2 Outcomes

We harmonized indicators of psychosocial health concerns from across the review form. Appendix A describes outcome definition rules and harmonization methods. We classified indicators into four outcome categories, plus an aggregate indicator of any psychosocial concern: 1) emotional, behavioral, and/or attentional (EBA); 2) personal and/or social; 3) speech, language, and/or communication (SLC); and 4) other developmental concerns. DRRB classified relevant Read v2 medical codes into these categories, with all classifications verified by a medical doctor (YAS) and disagreements resolved through discussion. We included indicators relevant to ongoing health, development, or wellbeing, and excluded acute and/or trivial issues. Each indicator was assigned a likelihood of *definite*, *probable*, or *possible*. This reflected the certainty that the recorded information indicated a concern in the relevant outcome. Example classifications, full code lists, outcome mappings, and likelihood mappings are provided in Tables A7–A10. For each child and outcome, we retained the highest indicator likelihood.

#### 2.3.3 Covariates

Adjusted analyses included sex, age at review in weeks, ethnicity, and area-level socioeconomic deprivation because of their established associations with preschool psychosocial outcomes [2, 16, 33, 39–46]. We derived ethnicity primarily from health review data using standard Public Health Scotland ethnicity codes [47], and used maternal ethnicity from the Scottish Birth Record where health review ethnicity was missing. Values coded as refused/not provided or not known were treated as missing. Known ethnicity was collapsed to *White* and *other ethnic group* because few children with known ethnicity were not White (9.6%). We measured deprivation using SIMD ranks grouped into five ordered, population-weighted deprivation groups, from most to least deprived, each containing approximately 20% of Scotland’s all-age population.

### 2.4 Statistical Analysis and Reporting

Primary analyses defined “likely cases” as children with any *definite* or *probable* outcome indicator. Sensitivity analyses defined “all potential cases” as children with any *definite*, *probable*, or *possible* indicator. We summarized cohort characteristics by care status and compared groups using Pearson’s chi-square tests for categorical variables and Welch’s *t*-test for age. We calculated unadjusted relative risks (RRs) and 95% confidence intervals (CIs) as ratios of group-specific prevalences [48, 49]. We estimated associations between care status and outcomes using logistic regression. For each outcome, we fitted three models: (1) unadjusted; (2) adjusted for age at review, sex, ethnicity, and area-level deprivation; and (3) adjusted with a care status by deprivation group interaction. Complete-case analyses included observations with complete data for sex, age, deprivation, and ethnicity. Absence of a recorded outcome indicator was treated as absence of that outcome. Analyses used R v4.5 [50] and Tidyverse packages [51]. This study is reported in accordance with the RECORD statement; a completed RECORD checklist is available as supplementary material [52, 53].

## 3 Results

### 3.1 Cohort Characteristics

Of the initial 488 702 observations available, 485 584 (99.4%) were eligible for inclusion by reported age. Of eligible records, 459 464 children (94.6%) had known care status, and 477 164 children (98.3%, including those with unknown care status) had complete covariate data. Ethnicity was the dominant source of missing covariate data, accounting for 8089 (96.1%) exclusions, with similar proportions in all care groups. Children with unknown care status were more likely to also have missing covariate data than children with known care status. The analytical cohort included 7887 children in care, 445 547 children not in care, and 23 730 children with unknown care status. Table 1 shows cohort characteristics stratified by care status. Compared with children not in care, children in care were more likely to live in deprived areas and less likely to be White. Children with unknown care status were less likely to live in the most deprived areas than both other care groups. This appears to be due to these children mainly living in health administrative regions (NHS Scotland Health Boards) of lower average deprivation and relates to systematic under-recording of demographic data in those regions. Sex distributions were similar across care status groups. Children in care were slightly younger at review, although the age difference was small.

**Table 1:**
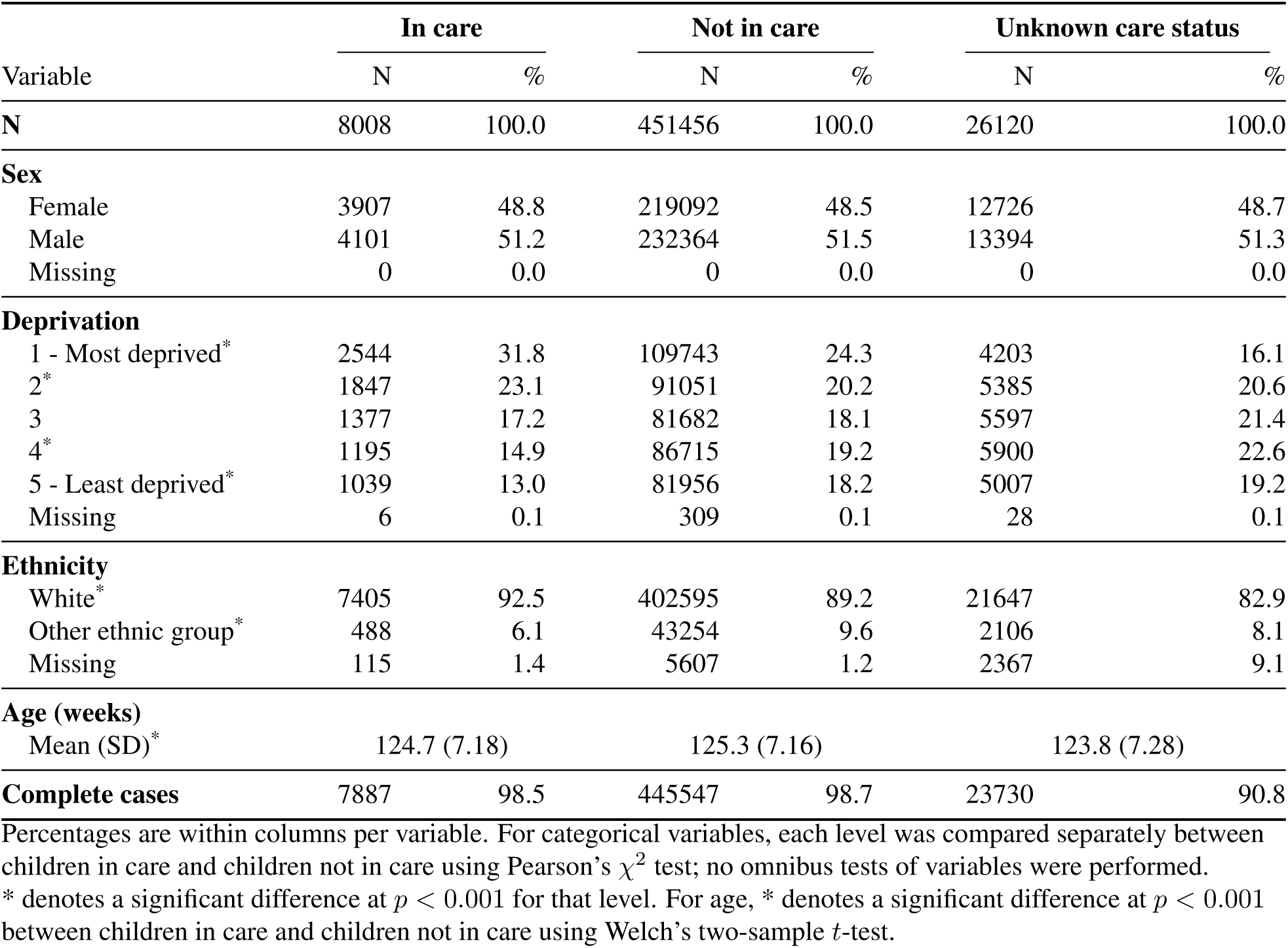
Cohort characteristics stratified by care status.

### 3.2 RQ1 - Differences in Psychosocial Health by Care Status

Of 7887 children in care, 2290 had at least one psychosocial concern (29.0%, 95% CI 28.0–30.0) compared with 77 836 of 445 547 children not in care (17.5%, 95% CI 17.4–17.6). Children in care had higher prevalence of all outcomes. Table 2 shows the prevalence for each separate outcome. Sensitivity analyses yielded similar findings (Table A11). Children with unknown care status had lower prevalence for all outcomes than both children in care and children not in care (see Tables A12–A15), possibly due to under-reporting of all variables in this group. Children in care were more likely than children not in care to have concerns recorded in one outcome category (16.0% vs 11.3%) and in two or more outcome categories (13.1% vs 6.2%).

**Table 2:**
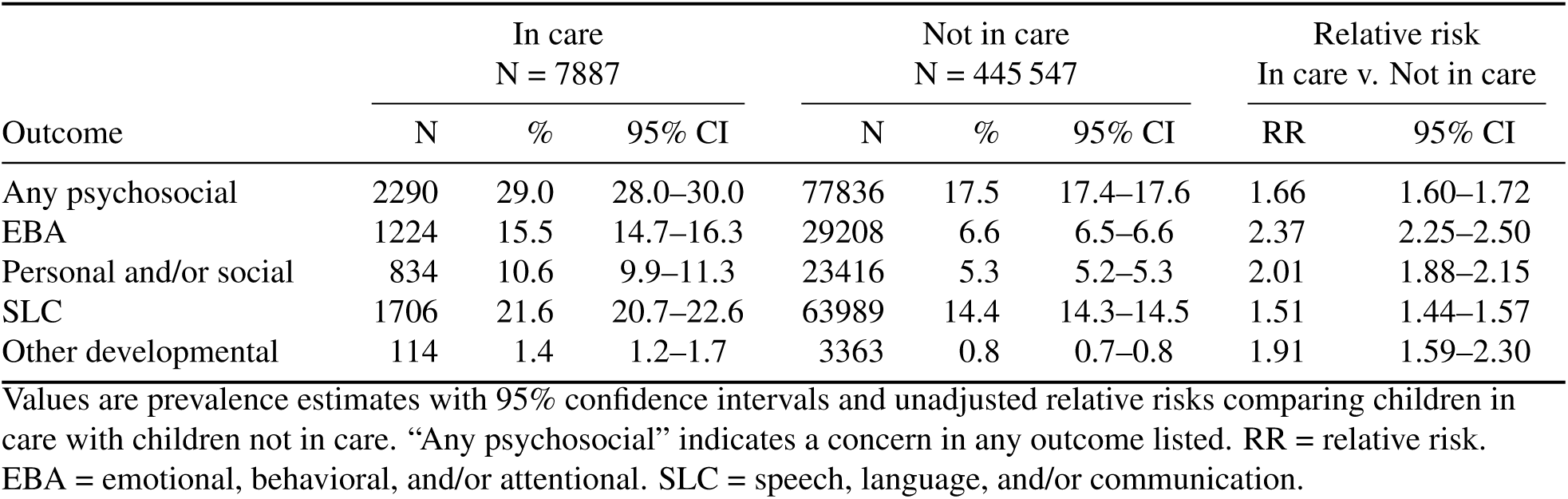
Prevalence and relative risks for psychosocial concerns by care status.

### 3.3 RQ2 - Care-Status Differences in Psychosocial Health by Deprivation

Figure 1 shows the prevalence of any psychosocial health concern by deprivation group and care status. For children in care the absolute difference in prevalence between the most and least deprived fifth of areas was 10.6 percentage points. For children not in care the absolute difference was greater at 14.2 percentage points. Sensitivity analyses showed similar trends (Figure A1), as did analysis of each individual outcome (Figures A2–A5).

**Figure 1:**
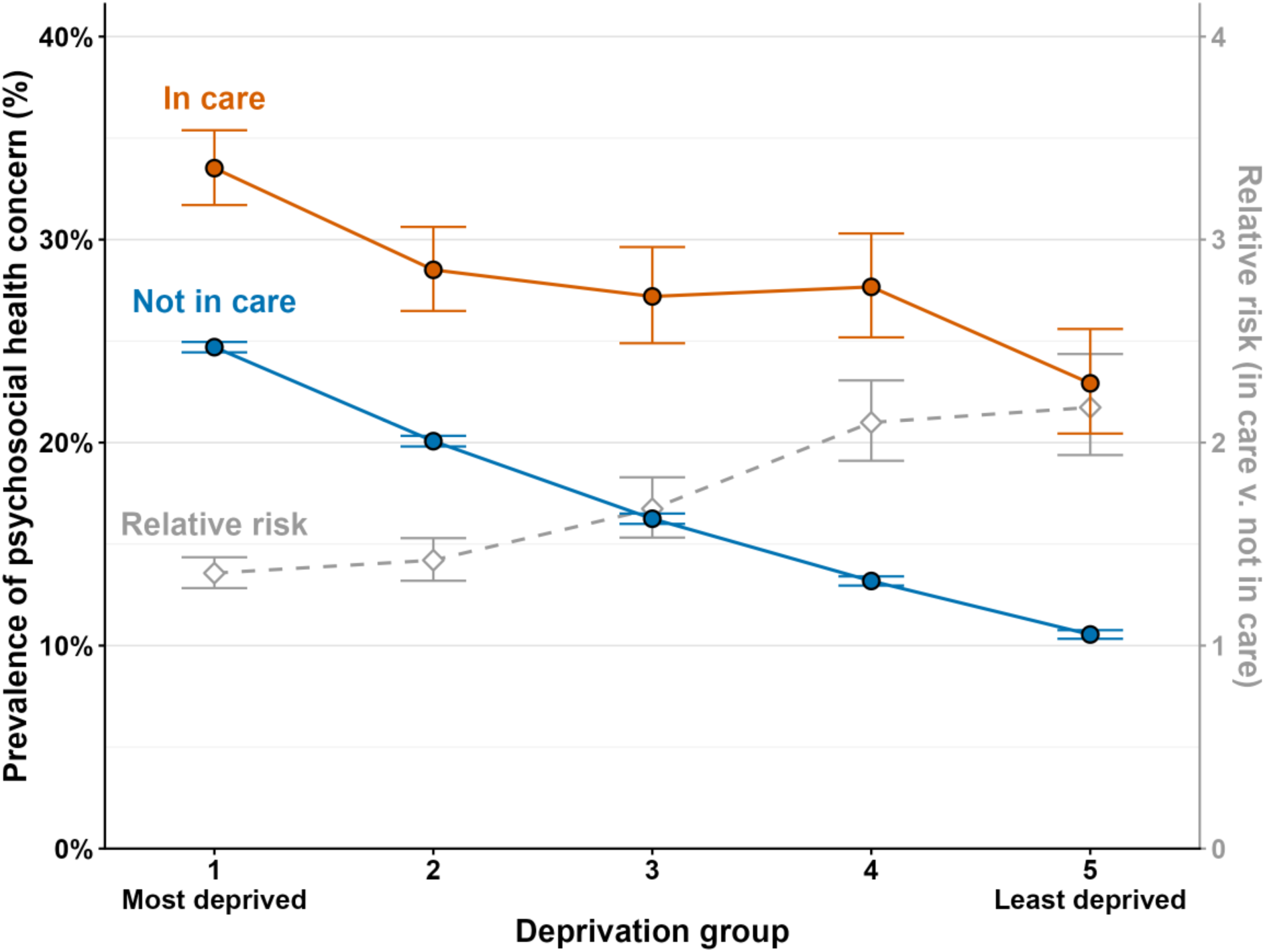
Prevalence of any psychosocial health concern by deprivation group and care status, with relative risks comparing children in care with children not in care. Circular points show prevalence estimates and 95% confidence intervals. Diamond points and the dashed line show unadjusted relative risks and 95% confidence intervals comparing children in care with children not in care within each deprivation group, using the right-hand vertical axis.

Figure 2 shows odds ratios comparing children in care with children not in care for each outcome. Across outcomes, odds ratios changed little after adjustment for covariates. Models including the care status by deprivation group interaction showed the odds ratio between children in care and not in care varied across deprivation groups. Odds ratios increased from more deprived areas to less deprived areas. Sensitivity analyses showed similar patterns (Figure A6). Supplementary model-related results are reported in Tables A16–A20.

**Figure 2:**
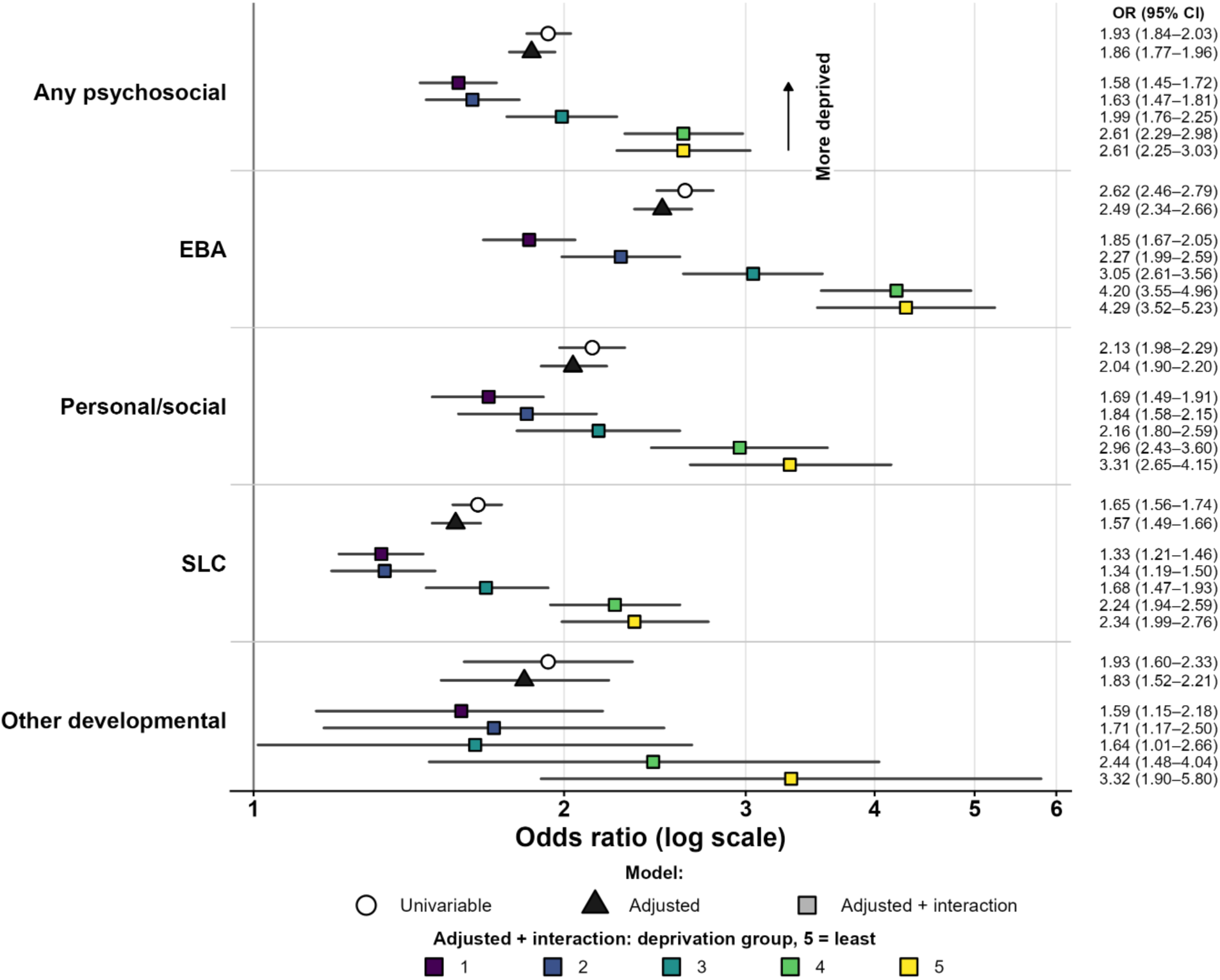
Odds ratios comparing children in care with children not in care across psychosocial outcomes. Points show odds ratios and lines show 95% confidence intervals. The figure shows estimates from (1) unadjusted models, (2) adjusted models without interaction, and (3) adjusted models including a care status by deprivation group interaction. For model (3), estimates are odds ratios for children in care compared with children not in care within each deprivation group. Deprivation group 1 is the most deprived fifth of areas and group 5 is the least deprived fifth. EBA = emotional, behavioral, and/or attentional. SLC = speech, language, and/or communication.

### 3.4 RQ3 - Deprivation Gradients in Psychosocial Health by Care Status

Prevalence of psychosocial health concerns increased with deprivation in both care groups. Table 3 reports unadjusted RRs and adjusted odds ratios. The table is stratified by care status and compares the most deprived with the least deprived fifth of areas. Results for intermediate deprivation groups are presented in Tables A26–A30. The deprivation gradient was larger for children not in care than children in care for all outcomes and measures. The proportion of children with concerns recorded for two or more individual outcomes also increased with deprivation in both groups; the absolute increase between the least and most deprived fifths was slightly larger for children not in care (3.1% to 9.5%) than children in care (10.6% to 14.9%).

**Table 3:** Relative risks and adjusted odds ratios comparing the most deprived with the least deprived fifth of areas, stratified by care status.

| Outcome | In care |  | Not in care |  |
| --- | --- | --- | --- | --- |
|  | RR (95% CI) | aOR (95% CI) | RR (95% CI) | aOR (95% CI) |
| Any psychosocial | 1.46 (1.29–1.66) | 1.71 (1.44–2.03) | 2.34 (2.29–2.40) | 2.82 (2.75–2.90) |
| EBA | 1.57 (1.30–1.90) | 1.65 (1.32–2.05) | 3.59 (3.45–3.75) | 3.83 (3.66–4.00) |
| Personal and/or social | 1.41 (1.12–1.76) | 1.46 (1.14–1.88) | 2.71 (2.59–2.84) | 2.87 (2.74–3.01) |
| SLC | 1.35 (1.17–1.57) | 1.49 (1.24–1.80) | 2.26 (2.20–2.32) | 2.63 (2.56–2.71) |
| Other developmental | 1.25 (0.67–2.32) | 1.26 (0.67–2.38) | 2.60 (2.30–2.95) | 2.64 (2.33–3.00) |

Estimates compare the most deprived fifth of areas with the least deprived fifth within each care-status group. Relative risks are unadjusted. Adjusted odds ratios are from the model including the care status by deprivation group interaction and adjustment for age at review, sex, and ethnicity. RR = relative risk. aOR = adjusted odds ratio. EBA = emotional, behavioral, and/or attentional. SLC = speech, language, and/or communication.

## 4 Discussion

This study provides population-level evidence that psychosocial health concerns are more common among two-year-old children in care than children not in care in Scotland. We also found evidence that the association between care status and psychosocial concern varied by deprivation group, with adjusted odds ratios decreasing as deprivation increased. Further, prevalence of psychosocial health concerns increased with deprivation in both care groups, but the socioeconomic gradient was steeper among children not in care. These findings suggest that the relative disadvantage to psychosocial health associated with being in care is attenuated in areas of greater deprivation. However, the core result was that prevalence was highest among children in care living in the most deprived fifth of areas for all outcomes and case definitions and results.

Our findings are consistent with prior evidence that psychosocial and developmental health concerns are more common among young children in care than children not in care [2, 4, 26]. For SLC concerns, the deprivation gradient among children not in care closely matched prior research in NHS Lothian, one of 14 health administrative regions in Scotland, using the same health review data and area-level deprivation measure [33]. Prevalence of EBA and personal and/or social concerns among children not in care also broadly matched other population-level analyses using the same dataset over a shorter period [34]. These comparisons support the face validity of our outcome definitions.

The difference in prevalence of psychosocial health problems between those in care and those not in care was greater in less deprived areas. For example, adjusted odds ratios for having at least one psychosocial concern ranged from 1.58 (95% CI 1.45–1.72) in the most deprived fifth of areas to 2.61 (95% CI 2.25–3.03) in the least deprived fifth. One explanation is that area-level deprivation may not capture household-level adversities associated with both entry to care and psychosocial development, such as parental mental health difficulties or problematic substance use. Children in care may therefore have elevated household-level risk across all area-level deprivation groups, making area-level deprivation a weaker marker of risk among children in care than among children not in care. This interpretation is supported by linked-data studies in the UK and Canada showing that household poverty, parental risk factors, and neighborhood poverty are each associated with entry to care, with household-level risks contributing information beyond area-level deprivation [16, 20, 54]. It is also consistent with a UK scoping review finding that individual-level measures of disadvantage tended to show stronger associations with early child health than area-level measures [55]. A scale issue is also possible. As prevalence of psychosocial health concerns rises among children not in care, *relative* measures comparing children in care with children not in care can narrow, even when *absolute* differences persist. A further possibility is that clinicians carrying out the health review may be more cautious when a child is known to be in care and use a lower threshold for recording concerns. For these reasons, variation in the strength of the care-status association across deprivation groups should be interpreted cautiously. The data show differential patterning by care status and deprivation, but do not identify the household, service, or placement mechanisms producing these differences. The key policy-relevant finding remains that absolute prevalence was highest among children in care living in the most deprived areas.

Of the four outcomes analyzed, EBA concerns showed the largest care-status differences and the greatest variation in care-status adjusted odds ratios across deprivation groups (ranging from 4.29 [95% CI 3.52–5.23] in the least deprived fifth to 1.85 [95% CI 1.67–2.05] in the most deprived fifth). This aligns with evidence that behavioral and externalizing problems are more common among children in foster care than children not in care, including preschool-aged children [3, 13]. However, externalizing problems among children in foster care vary by maltreatment type and adversity history [56]. Future studies should include information on care trajectories where feasible.

### 4.1 Strengths

We used near-universal health review data to compare psychosocial concerns in two-year-old children in care and children not in care. This addresses limitations in existing studies, which often lack comparator groups, include broad age ranges, or use data collected at entry to care which may be a point of acute health need [28, 57]. The study period provided a large population-based sample. We harmonized outcomes across all 27–30 Month Health Review fields, reducing reliance on a single limited section. Using two case definitions also showed that findings were robust to broader outcome classification.

### 4.2 Limitations

Because data were cross-sectional, we could not infer causal mechanisms. Data were secondary, so recorded concerns may reflect recording practices as well as underlying psychosocial health need. Deprivation was measured at area level rather than household level, which may misclassify household circumstances and attenuate socioeconomic gradients [58]. Several potentially important confounders were unavailable at sufficiently low levels of missingness, including gestational age, birthweight, and disability [18, 33, 59–63]. We aggregated all placement types into a single care category and lacked placement history data, so children previously, but not currently, in care were included as children not in care. Public Health Scotland noted that care status may also have been over-recorded in the early years of the health review data, where some children recorded as “looked after at home” may have simply been living at home with no special health or social service involvement. Any over-recording would bias associations towards the null. We manually classified health review indicators into four outcome categories, which introduced some subjectivity, though we applied the same mapping by care status and a medical doctor (YAS) verified the classification. Findings are most directly applicable to Scotland and may not generalize to settings with different care systems, early-years health services, or policy contexts.

### 4.3 Conclusion

Psychosocial concerns were more common among preschool-aged children in care than children not in care. Prevalence increased with area-level deprivation in both groups, and children in care living in the most deprived areas had the highest burden. These findings support timely identification and support for children in care, alongside broader policies to reduce socioeconomic deprivation.

## Supporting information

RECORD guideline

## Data Availability

Data are available via application to the NHS Scotland Public Benefit and Privacy Panel for Health and Social Care.

## Declarations

### Data availability

The data used in this study are not publicly available because they contain sensitive health records. Researchers seeking access to these data can apply through the relevant Public Health Scotland data governance processes, referring where appropriate to Public Benefit and Privacy Panel–Health and Social Care approval reference 2021-0199.

### Code availability

Analysis code is available from the corresponding author on reasonable request.

### Author contributions

DRRB contributed to conceptualization, data curation, formal analysis, investigation, methodology, resources, software, visualization, writing the original draft, and writing review and editing. DB, ADMcM, and MA contributed to conceptualization, methodology, resources, supervision, and writing review and editing. AHL contributed to methodology, supervision, and writing review and editing. YAS contributed to methodology and writing review and editing.

## Acknowledgments

Thank you to the eDRIS Team at Public Health Scotland for their support in obtaining approvals, provisioning and linking data, and the use of the secure analytical platform within the National Safe Haven. Particularly, we thank James Watson, eDRIS Principal Information Development Manager, for his assistance and support throughout this work.

## References

[1] K. Dubois-Comtois et al. “Are children and adolescents in foster care at greater risk of mental health problems than their counterparts? A *meta*-analysis”. In: Children and Youth Services Review 127 (2021), p. 106100. DOI: 10.1016/j.childyouth.2021.106100.

[2] J. D. McLeigh, K. Tunnell, and C. Lazcano. “Developmental Status of Young Children in Foster Care”. In: Journal of Developmental & Behavioral Pediatrics 42.5 (2021), p. 389. DOI: 10.1097/DBP.0000000000000906.

[3] K. Turney and C. Wildeman. “Mental and Physical Health of Children in Foster Care”. In: Pediatrics 138.5 (2016), e20161118. DOI: 10.1542/peds.2016-1118.

[4] A. D. Engler et al. “A Systematic Review of Mental Health Disorders of Children in Foster Care”. In: Trauma, Violence, & Abuse 23.1 (2022), pp. 255–264. DOI: 10.1177/1524838020941197.

[5] T. Ford et al. “Psychiatric disorder among British children looked after by local authorities: Comparison with children living in private households”. In: British Journal of Psychiatry 190.4 (2007), pp. 319–325. DOI: 10.1192/bjp.bp.106.025023.

[6] N. Hu et al. “Developmental trajectories of socio-emotional outcomes of children and young people in out-of-home care – Insights from data of Pathways of Care Longitudinal Study (POCLS)”. In: Child Abuse & Neglect 149 (2024), p. 106196. DOI: 10.1016/j.chiabu.2023.106196.

[7] L. K. Leslie et al. “The Physical, Developmental, and Mental Health Needs of Young Children in Child Welfare by Initial Placement Type”. In: Journal of Developmental & Behavioral Pediatrics 26.3 (2005), pp. 177–185. DOI:10.1097/00004703-200506000-00003.

[8] E. Fernandez. “Reasons for Children Entering Care”. In: Accomplishing Permanency: Reunification Pathways and Outcomes for Foster Children. Ed. by E. Fernandez. Dordrecht: Springer Netherlands, 2013, pp. 31–44. ISBN: 978-94-007-5092-0. DOI:10.1007/978-94-007-5092-0_3.

[9] E. Neil, L. Gitsels, and J. Thoburn. “Children in care: Where do children entering care at different ages end up? An analysis of local authority administrative data”. In: Children and Youth Services Review 106 (2019), p. 104472. DOI: 10.1016/j.childyouth.2019.104472.

[10] Welsh Government. Children looked after by local authorities, April 2023 to March 2024. Cardiff: Welsh Government, 2025. URL: https://www.gov.wales/children-looked-after-local-authorities-april-2023-march-2024.

[11] Office for National Statistics. Who are the children entering care in England? Office for National Statistics. 2022. URL: https://www.ons.gov.uk/peoplepopulationandcommunity/healthandsocialcare/socialcare/articles/whoarethechildrenenteringcareinengland/2022-11-04.

[12] E. Lee et al. “The Cumulative Effect of Prior Maltreatment on Emotional and Physical Health of Children in Informal Kinship Care”. In: Journal of Developmental & Behavioral Pediatrics 41.4 (2020), pp. 299–307. DOI: 10.1097/DBP.0000000000000769.

[13] S. R. Horn et al. “Polyvictimization and externalizing symptoms in foster care children: The moderating role of executive function”. In: Journal of Trauma & Dissociation 19.3 (2018), pp. 307–324. DOI: 10.1080/15299732.2018.1441353.

[14] J. Yu, D. L. Haynie, and S. E. Gilman. “Patterns of Adverse Childhood Experiences and Neurocognitive Development”. In: JAMA Pediatrics 178.7 (2024), pp. 678–687. DOI: 10.1001/jamapediatrics.2024.1318.

[15] A. D. McMahon et al. “Inequalities in the dental health needs and access to dental services among looked after children in Scotland: A population data linkage study”. In: Archives of Disease in Childhood 103.1 (2018), pp. 39–43. DOI:10.1136/archdischild-2016-312389.

[16] L. L. Roos, E. Wall-Wieler, and J. B. Lee. “Poverty and Early Childhood Outcomes”. In: Pediatrics 143.6 (2019), e20183426. DOI: 10.1542/peds.2018-3426.

[17] G. C. M. Skinner, N. Hodges, and E. Kennedy. “A Systematic Review of the Relationship between Economic Inequalities, the Social Gradient and Child Abuse and Neglect”. In: Child & Youth Services 46.4 (2025), pp. 811–869. DOI: 10.1080/0145935X.2025.2456625.

[18] D. Brown et al. “Mortality outcomes of children and young people who have spent time in care: evidence from Children’s Health in Care in Scotland, a population-wide administrative data cohort study”. In: Archives of Disease in Childhood 110.10 (2025), pp. 837–843. DOI: 10.1136/archdischild-2024-327854.

[19] P. Bywaters. “Inequalities in child welfare: Towards a new policy, research and action agenda”. In: British Journal of Social Work 45.1 (2015), pp. 6–23. DOI: 10.1093/bjsw/bct079.

[20] N. Warner et al. “What affects the likelihood of children entering public care? The interaction of household low income, area-level deprivation and parental risk factors”. In: Children and Youth Services Review 183 (2026), p. 108851. DOI: 10.1016/j.childyouth.2026.108851.

[21] S. Goldfeld et al. “The impact of multidimensional disadvantage over childhood on developmental outcomes in Australia”. In: International Journal of Epidemiology 47.5 (2018), pp. 1485–1496. DOI: 10.1093/ije/dyy087.

[22] N. L. Hair et al. “Association of Child Poverty, Brain Development, and Academic Achievement”. In: JAMA pediatrics 169.9 (2015), pp. 822–829. DOI: 10.1001/jamapediatrics.2015.1475.

[23] S. A. Rosenberg, D. Zhang, and C. C. Robinson. “Prevalence of developmental delays and participation in early intervention services for young children”. In: Pediatrics 121.6 (2008), e1503–1509. DOI: 10.1542/peds.2007-1680.

[24] F. Lima et al. “Infants entering out-of-home care: Health, developmental needs and service provision”. In: Child Abuse & Neglect 149 (2024), p. 106577. DOI: 10.1016/j.chiabu.2023.106577.

[25] L. de Montaigne et al. “Étude des notifications à la Maison départementale des personnes handicapées chez les enfants placés à l’Aide sociale à l’enfance dans les Bouches-du-Rhône”. In: Archives de Pédiatrie 22.9 (2015), pp. 932–942. DOI: 10.1016/j.arcped.2015.06.018.

[26] M. Vasileva and F. Petermann. “Attachment, Development, and Mental Health in Abused and Neglected Preschool Children in Foster Care: A Meta-Analysis”. In: Trauma, Violence, and Abuse 19.4 (2018), pp. 443–458. DOI: 10.1177/1524838016669503.

[27] J. C. Krier, T. D. Green, and A. Kruger. “Youths in foster care with language delays: Prevalence, causes, and interventions”. In: Psychology in the Schools 55.5 (2018), pp. 523–538. DOI: 10.1002/pits.22129.

[28] D. R. R. Bradford et al. Physical health of care-experienced young children in high-income countries: A scoping review. 2025. DOI: 10.1101/2025.04.15.25325761. Pre-published.

[29] T. Hillen et al. “Assessing the prevalence of mental health disorders and mental health needs among preschool children in care in England”. In: Infant Mental Health Journal 33.4 (2012), pp. 411–420. DOI: 10.1002/imhj.21327.

[30] S. Penttilä, et al. “Child- and parent-related determinants for out-of-home care in a nationwide population with neurodevelopmental disorders: a register-based Finnish birth cohort 1997 study”. In: European Child & Adolescent Psychiatry 33.10 (2024), pp. 3459–3470. DOI: 10.1007/s00787-024-02406-w.

[31] Scottish Government. Universal Health Visiting Pathway in Scotland: pre-birth to pre-school. 2015. URL: https://www.gov.scot/publications/universal-health-visiting-pathway-scotland-pre-birth-pre-school/.

[32] Public Health Scotland. Child health pre-school review coverage 2023 to 2024. 2025. URL: https://publichealthscotland.scot/publications/child-health-pre-school-review-coverage/child-health-pre-school-review-coverage-2023-to-2024/ (visited on 09/03/2025).

[33] D. Ene et al. “Associations of Socioeconomic Deprivation and Preterm Birth with Speech, Language, and Communication Concerns among Children Aged 27 to 30 Months”. In: JAMA Network Open 2.9 (2019), pp. 1–11. DOI:10.1001/jamanetworkopen.2019.11027.

[34] I. Hardie et al. “COVID-19 public health and social measures (PHSM) and early childhood developmental concerns in Scotland: an interrupted time series analysis”. In: The Lancet Regional Health - Europe 60 (2026), p. 101525. DOI: 10.1016/j.lanepe.2025.101525.

[35] Public Health Scotland. Child health pre-school review coverage: 2024 to 2025. 2026. URL: https://www.publichealthscotland.scot/publications/child-health-pre-school-review-coverage/child-health-pre-school-review-coverage-2024-to-2025/ (visited on 05/03/2026).

[36] D. R. R. Bradford et al. “Assessing the risk of endogeneity bias in health and mortality inequalities research using composite measures of multiple deprivation which include health-related indicators: A case study using the Scottish Index of Multiple Deprivation and population health and mortality data”. In: Health & Place 80 (2023), p. 102998. DOI: 10.1016/j.healthplace.2023.102998.

[37] G. McCartney et al. “How important is it to avoid indices of deprivation that include health variables in analyses of health inequalities?” In: Public Health 221 (2023), pp. 175–180. DOI: 10.1016/j.puhe.2023.06.028.

[38] Scottish Government. SIMD 2020 technical notes. Edinburgh: Scottish Government, 2020. URL: https://www.gov.scot/publications/simd-2020-technical-notes/.

[39] A. Demirci and M. Kartal. “The prevalence of developmental delay among children aged 3-60months in Izmir, Turkey”. In: Child: Care, Health and Development 42.2 (2016), pp. 213–219. DOI: 10.1111/cch.12289.

[40] G. Pratte et al. “Participation in Activities Fostering Children’s Development and Parental Concerns about Children’s Development: Results from a Population-Health Survey of Children Aged 0–5 Years in Quebec, Canada”. In: International Journal of Environmental Research and Public Health 17.8 (2020), p. 2878. DOI: 10.3390/ijerph17082878.

[41] N. Razaz et al. “Five-minute Apgar score as a marker for developmental vulnerability at 5 years of age”. In: Archives of Disease in Childhood - Fetal and Neonatal Edition 101.2 (2016), F114–F120. DOI: 10.1136/archdischild-2015-308458.

[42] S. J. Barry et al. “Mapping area variability in social and behavioural difficulties among Glasgow pre-schoolers: Linkage of a survey of pre-school staff with routine monitoring data”. In: Child: Care, Health and Development 41.6 (2015), pp. 853–864. DOI: 10.1111/cch.12237.

[43] F. Sim et al. “Preschool developmental concerns and adjustment in the early school years: Evidence from a Scottish birth cohort”. In: Child: Care, Health and Development 45.5 (2019), pp. 719–736. DOI: 10.1111/cch.12695.

[44] M. Curtin et al. “Determinants of vulnerability in early childhood development in Ireland: a cross-sectional study”. In: BMJ Open 3.5 (2013), e002387. DOI: 10.1136/bmjopen-2012-002387.

[45] W. D. Lohr et al. “Antipsychotic Medications for Low-Income Preschoolers: Long Duration and Psychotropic Medication Polypharmacy”. In: Psychiatric Services 73.5 (2022), pp. 510–517. DOI: 10.1176/appi.ps.202000673.

[46] M. J. Childs et al. “Disabilities in children receiving social care and support in Wales and factors associated with placement into care: A population-based data linkage study”. In: Child Abuse & Neglect 166 (2025), p. 107510. DOI: 10.1016/j.chiabu.2025.107510.

[47] Public Health Scotland. Data Dictionary - Ethnic Group. 2023. URL: https://publichealthscotland.scot/resources-and-tools/health-intelligence-and-data-management/national-data-catalogue/data-dictionary/search-the-data-dictionary/ethnic-group/ (visited on 06/01/2026).

[48] B. M. Katz. “Tests for Equality of Correlated Proportions in a Polychotomous Response Design”. In: Journal of Educational Statistics 3.2 (1978), pp. 145–178. DOI: 10.3102/10769986003002145.

[49] M. W. Fagerland, S. Lydersen, and P. Laake. “Recommended tests and confidence intervals for paired binomial proportions”. In: Statistics in Medicine 33.16 (2014), pp. 2850–2875. DOI: 10.1002/sim.6148.

[50] R Core Team. R: A Language and Environment for Statistical Computing. Vienna, Austria, 2022. URL: https://www.r-project.org.

[51] H. Wickham et al. “Welcome to the {tidyverse}”. In: J. Open Source Softw. 4.43 (2019), p. 1686. DOI: 10.21105/joss.01686.

[52] E. I. Benchimol et al. “The REporting of studies Conducted using Observational Routinely-collected health Data (RECORD) Statement”. In: PLOS Medicine 12.10 (2015). Ed. by Y.-K. Tu, e1001885. DOI: 10.1371/journal.pmed.1001885.

[53] J. P. Vandenbroucke et al. “Strengthening the Reporting of Observational Studies in Epidemiology (STROBE): Explanation and Elaboration”. In: PLOS Medicine 4.10 (2007), e297. DOI: 10.1371/journal.pmed.0040297.

[54] N. Warner et al. “Parental risk factors and children entering out-of-home care: The effects of cumulative risk and parent’s sex”. In: Children and Youth Services Review 160 (2024), p. 107548. DOI: 10.1016/j.childyouth.2024.107548.

[55] A. Clery et al. “Measuring disadvantage in the early years in the UK: A systematic scoping review”. In: SSM - Population Health 19 (June 2022), p. 101206. DOI: 10.1016/j.ssmph.2022.101206.

[56] K. C. Pears, H. K. Kim, and P. A. Fisher. “Psychosocial and cognitive functioning of children with specific profiles of maltreatment”. In: Child Abuse & Neglect 32.10 (2008), pp. 958–971. DOI: 10.1016/j.chiabu.2007.12.009.

[57] G. D. Batty, M. Kivimäki, and P. Frank. “State care in childhood and adult mortality: a systematic review and meta-analysis of prospective cohort studies”. In: The Lancet Public Health 7.6 (2022), e504–e514. DOI: 10.1016/S2468-2667(22)00081-0.

[58] G. McCartney et al. “How well do area-based deprivation indices identify income- and employment-deprived individuals across Great Britain today?” In: Public Health 217 (2023), pp. 22–25. DOI: 10.1016/j.puhe.2023.01.020.

[59] S. Nivins et al. “Gestational Age and Cognitive Development in Childhood”. In: JAMA Network Open 8.4 (2025), e254580. DOI: 10.1001/jamanetworkopen.2025.4580.

[60] J. L. Cheong et al. “Association Between Moderate and Late Preterm Birth and Neurodevelopment and Social-Emotional Development at Age 2 Years”. In: JAMA Pediatrics 171.4 (2017), e164805. DOI: 10.1001/jamapediatrics.2016.4805.

[61] J. L. Y. Cheong et al. “Neurodevelopment at Age 9 Years Among Children Born at 32 to 36 Weeks’ Gestation”. In: JAMA network open 7.11 (2024), e2445629. DOI: 10.1001/jamanetworkopen.2024.45629.

[62] J. Burns et al. “Comparing outcomes of children and youth with fetal alcohol spectrum disorder (FASD) in the child welfare system to those in other living situations in Canada: Results from the Canadian National FASD Database”. In: *Child: Care*, Health and Development 47.1 (2021), pp. 77–84. DOI: 10.1111/cch.12817.

[63] L. Shea et al. “Foster Care Involvement Among Youth With Intellectual and Developmental Disabilities”. In: JAMA Pediatrics 178.4 (2024), pp. 384–390. DOI: 10.1001/jamapediatrics.2023.6580.

[64] J. Squires et al. ASQ-3 Technical Report. Paul H. Brookes Publishing Co., 2009. URL: https://agesandstages.com/wp-content/uploads/2025/06/ASQ-3-Technical-Appendix_web_2025.pdf.

