## Appendices for "Psychosocial Health Inequalities and Socioeconomic Deprivation Gradients Among Preschool Children in Care and Not in Care: An Administrative Health Data Study"

### Appendix A Outcome Classification Rules

The 27–30 Month Health Review form records information in structured sections. This appendix describes how we used the information in these sections to derive whether each child had each of the four outcome categories used in this study. We mapped eligible recorded information to one or more outcome categories and assigned a likelihood rating of *definite*, *probable*, or *possible*. We counted a child as having an outcome only if at least one source item met a mapping rule for that outcome. When a source item was blank, unavailable in that form version, recorded as incomplete, or recorded as information not recorded, we treated it as providing no evidence of that outcome. We counted the child as not having that outcome unless another source item met a mapping rule.

#### Appendix A.1 Structured Developmental Concern Fields

The developmental section of the 27–30 Month Health Review form records whether the caregiver and/or practitioner identified concerns in specific developmental domains. The form versions used during the study period differed slightly, with revised developmental fields introduced in 2017. Earlier form versions recorded separate emotional, behavioral, attention, social, and speech, language, and communication domains. Later form versions combined some domains, e.g. separate fields for emotional, behavioral, and attentional concerns were combined into a single composite field for all three. We harmonized fields from both versions where they captured comparable developmental domains. Table A1 shows how we mapped form domains to outcome categories.

Each developmental field could record no concern, a newly suspected concern, a previously identified concern or disorder, an incomplete assessment, or information not recorded. Table A2 shows how we mapped these response levels to likelihood ratings.

Table A1: Mapping of structured developmental form domains to outcome categories.

| **Form domain** | **Outcome category** |
|---|---|
| Emotional (Pre-2017) | Emotional, behavioral, and/or attentional |
| Behavioral (Pre-2017) | Emotional, behavioral, and/or attentional |
| Attention (Pre-2017) | Emotional, behavioral, and/or attentional |
| Emotional/Behavioral | Emotional, behavioral, and/or attentional |
| Social (Pre-2017) | Personal and/or social |
| Personal/Social | Personal and/or social |
| Speech, Language & Communication (Pre-2017) | Speech, language, and/or communication |
| Speech, Language & Communication | Speech, language, and/or communication |

Table A2: Mapping of structured developmental concern responses to likelihood ratings.

| **Form response** | **Likelihood assigned** |
|---|---|
| No Concerns | None assigned |
| Concern newly suspected | *Probable* |
| Concern/Disorder previously identified | *Probable* |
| Assessment incomplete | None assigned |
| Information not recorded | None assigned |

#### Appendix A.2 Future Action and Referral Fields

The 27–30 Month Health Review form includes future action fields that record whether the practitioner documented further action for specific developmental domains. These fields could record actions such as support being provided, requested, discussed, signposted, or refused. For this study, only the Speech, Language & Communication future action domain contributed to the outcomes. We treated responses indicating provided or requested action as *probable* evidence of a speech, language, and/or communication concern. We treated discussed, signposted, or refused action as *possible* evidence of a speech, language, and/or communication concern. Table A3 shows this mapping.

Table A3: Mapping of Speech, Language & Communication future action responses to likelihood ratings.

| **Form response** | **Likelihood assigned** |
|---|---|
| Provided | *Probable* |
| Requested | *Probable* |
| Discussed | *Possible* |
| Signposted | *Possible* |
| Refused | *Possible* |

#### Appendix A.3 Parent/Carer Concern Fields

The 27–30 Month Health Review form includes a carer concern item which allows the practitioner to mark whether the carer raised concerns about feeding/diet, growth/weight, sleep, development, physical health, or another concern recorded in free text. For this study, only a marked concern about development contributed to the outcomes. We treated this as a nonspecific developmental indicator and conservatively assigned it as *possible* for emotional, behavioral, and/or attentional concerns and personal and/or social concerns.

#### Appendix A.4 ASQ-3 Scores

The 27–30 Month Health Review form can, but infrequently did in our data, include scores from the Ages and Stages Questionnaire (Third Edition; ASQ-3). The ASQ-3 is an age-specific developmental screening tool, with different versions used at different stages of early childhood development. We presumed the ASQ-3 version using each child's age in completed weeks at review, based on the age ranges in the ASQ-3 technical manual [64]. Table A4 shows this mapping.

Table A4: Mapping of age at review to ASQ-3 version.

| **Age at review in completed weeks** | **ASQ-3 version assigned** |
|---|---|
| 104–109 | 24 months |
| 110–122 | 27 months |
| 123–135 | 30 months |
| 136–148 | 33 months |
| 149–156 | 36 months |

For this study, we mapped the communication domain of the ASQ-3 to our speech, language, and/or communication outcome and personal-social domain to our personal and/or social outcome. Table A5 reports the empirical cut-off scores extracted from the ASQ-3 technical manual [64]. For each ASQ-3 version and domain, these represent 1 and 2 standard deviations below the mean. We used scores at or below the 2 SD cut-off to indicate *probable* concern and scores at or below the 1 SD cut-off but above the 2 SD cut-off to indicate *possible* concern.

Table A5: ASQ-3 version-specific cut-off scores used to classify speech, language, and/or communication and personal and/or social categories.

| ASQ-3 version | Outcome category | Possible concern | Probable concern |
|---|---|---|---|
| 24 months | Speech, language, and/or communication | 38.70 | 25.17 |
| 24 months | Personal and/or social | 41.34 | 31.54 |
| 27 months | Speech, language, and/or communication | 37.22 | 24.02 |
| 27 months | Personal and/or social | 36.11 | 25.31 |
| 30 months | Speech, language, and/or communication | 43.56 | 33.30 |
| 30 months | Personal and/or social | 41.94 | 32.01 |
| 33 months | Speech, language, and/or communication | 37.37 | 25.36 |
| 33 months | Personal and/or social | 39.95 | 28.96 |
| 36 months | Speech, language, and/or communication | 41.43 | 30.99 |
| 36 months | Personal and/or social | 44.07 | 35.33 |

Possible concern corresponds to the 1 SD cut-off and probable concern corresponds to the 2 SD cut-off.

#### Appendix A.5 Read v2 Codes

Read v2 codes are part of a hierarchical clinical coding system used in UK primary and community health records. Codes with trailing full stops represent higher-level parent codes, with more specific child codes replacing full-stop placeholders with additional characters. We used Read v2 codes to supplement structured health review fields for care status and psychosocial or developmental concerns.

##### Appendix A.5.1 Care Status Codes

Table A6 lists the Read v2 codes indicating that a child was in care. These codes captured foster care, looked-after child status, kinship care, prospective adoption, and related care arrangements. We combined these codes with the structured care-status variable in the health review data. Only 42 children were classified as in care from Read v2 codes alone.

Table A6: Read v2 codes indicating a child is in care.

| Code | Description | Code | Description |
|---|---|---|---|
| 1338. | Fostered | 8GE81 | Child for adoption |
| 13IB. | Child in care | 918F3 | Has kinship carer |
| 13IB0 | Child in foster care | 918F4 | Lives with prospective adopter |
| 13IB1 | Looked after child | 13FH. | Lives with relatives |
| 13II. | Child deserted by parents | 13Ic. | Child lives with another relative |
| 13Iv. | Looked after child - Children (Scotland) Act 1995 | 13IK. | Child lives with grandparents |
| 13VJ. | In care | 13It. | Lives with grandmother |
| 38C0. | Child in care health assessment | 13Iu. | Child living with unrelated adult |
| 38C00 | Looked after child initial health assessment | 13Ix. | Child for permanence |
| 38C01 | Looked after child health assessment 6 month review | 8GEA. | Care from relatives |
| 8GE7. | Foster care | | |

##### Appendix A.5.2 Psychosocial and Developmental Concern Codes

We classified Read v2 codes indicating psychosocial or developmental concerns into the study outcome categories using the decision rules below. These rules guided code inclusion, outcome assignment, and likelihood rating.

- Each Read v2 code was classified into up to three outcome categories using the information available in the code description.

  *Example:* `1B1J0` "Behavioural, emotional and social difficulties" was classified as *definite* for behavioral, emotional, and social concerns.

- Codes were assigned to an outcome only where they were considered relevant to the child's ongoing health, development, or wellbeing. Trivial and minor acute conditions were excluded.

  *Example:* `F4C0.` "Acute conjunctivitis" was excluded.

- Each included code was assigned a likelihood category of *definite*, *probable*, or *possible*. This reflected the certainty that the recorded information indicated a concern in the relevant outcome category.

  *Example:* `E2F3.` "Speech or language developmental disorder" was classified as *definite*. `Ryu6.` "[X]Symptoms and signs involving speech and voice" was classified as *probable*. `75023` "Unilateral lip adhesion" was classified as *possible*.

- Codes indicating a named developmental disorder, established diagnosis, ongoing medication, specialized medical equipment use, or receipt of a specific therapy or treatment were classified as *definite*.

  *Example:* `E140.` "Infantile autism" was classified as *definite* for other developmental concerns. `9NNj2` "Under care of speech and language therapist" was classified as *definite* for speech, language, and/or communication concerns.

- Codes indicating referral to, signposting to, attendance at, review by, being seen by, or being under the care of a relevant specialist service or health professional were classified as *probable*, unless they indicated receipt of a specific therapy or treatment.

  *Example:* `9N0Q.` "Seen in speech and language clinic" was classified as *probable* for speech, language, and/or communication concerns.

- Codes described as "history of" were classified as *possible*, unless the relevant condition was clearly an ongoing concern.

  *Example:* `1469.` "H/O: behaviour problem" was classified as *possible* for behavioral concerns.

- Codes indicating operations or procedures on specific body parts were classified as *possible*. Exceptions were codes that referred only to an acute, transient, or resolved event, which were excluded.

  *Example:* `314..` "Special ENT procedures" was classified as *possible* for speech, language, and/or communication concerns.

- Codes indicating non-standard assessment were classified as *possible*. Non-standard assessment was defined as assessment outside universal components of the 27–30 Month Health Review.

  *Example:* `311B.` "Cognitive assessment" was classified as *possible* for other developmental concerns.

- Where code descriptions were ambiguous, subjective, or context dependent, they were typically assigned a *possible* likelihood.

  *Example:* `E273.` "Stereotyped repetitive movements" was classified as *possible* rather than *definite*.

- Perinatal and birth-related codes were excluded, except where the code clearly indicated an ongoing child health, developmental, or psychosocial concern at the age of review, such as a congenital condition.

  *Example:* `Q....` "Perinatal conditions" was excluded. `P83yX` "Congenital malformation of larynx, unspecified" contributed as a possible speech, language, and/or communication concern.

- Codes explicitly described as acute, transient, resolved, trivial, administrative, or uninformative were excluded unless the code also indicated an ongoing concern.

  *Example:* `H061.` "Acute bronchiolitis" was excluded.

- Codes for conditions judged not to be age appropriate for children aged approximately 27–30 months were excluded.

  *Example:* `Eu401` "[X]Social phobias" and `Eu50.` "[X]Eating disorders" were excluded.

##### Appendix A.5.3 Emotional, Behavioral, and/or Attentional

Table A7: Read v2 codes contributing to emotional, behavioral, and/or attentional concerns.

| **Code** | **Description** | **Likelihood** |
|---|---|---|
| 13Z4C | Behavioural problems at school | Possible |
| 1469. | H/O: behaviour problem | Possible |
| 1A22. | Enuresis | Possible |
| 1A220 | Nocturnal enuresis | Possible |
| 1A221 | Daytime enuresis | Possible |
| 1A23. | Incontinence of urine | Possible |
| 1B1J. | Emotional problem | Definite |
| 1B1J0 | Behavioural, emotional and social difficulties | Definite |
| 1B1J1 | Emotional behavioural difficulties | Definite |
| 1B1X. | Behavioural problem | Definite |
| 1B24. | Has a tic | Definite |
| 1BR.. | Reduced concentration | Possible |
| 1BR0. | Reduced concentration span | Possible |
| 1P00. | Hyperactive behaviour | Definite |
| 1P3.. | Compulsive behaviour | Probable |
| 28B.. | O/E - easily distractable | Possible |
| 3973. | Difficulty performing toileting activities | Possible |
| 3AB.. | Behaviour assessment | Possible |
| 8GL.. | Attachment-based therapy | Definite |
| E272. | Tics | Possible |
| E2720 | Tic disorder unspecified | Possible |
| E272z | Tic NOS | Possible |
| E273. | Stereotyped repetitive movements | Possible |
| E2752 | Pica | Definite |
| E27z0 | Hair plucking | Definite |
| E2920 | Separation anxiety disorder | Definite |
| E2C.. | Disturbance of conduct NEC | Definite |
| E2C1. | Nonaggressive unsocial conduct disorder | Possible |
| E2C2. | Socialised conduct disorder | Definite |
| E2Cz. | Unspecified disturbance of conduct | Definite |
| E2Czz | Disturbance of conduct NOS | Possible |
| E2D2. | Childhood and adolescent disturbance with sensitivity | Possible |
| E2D20 | Childhood and adolescent disturbance with shyness | Possible |
| E2E.. | Childhood hyperkinetic syndrome | Definite |
| E2E0. | Child attention deficit disorder | Definite |
| E2E00 | Attention deficit without hyperactivity | Definite |
| E2E01 | Attention deficit with hyperactivity | Definite |
| Eu056 | [X]Organic emotionally labile [asthenic] disorder | Definite |
| Eu42. | [X]Obsessive - compulsive disorder | Definite |
| Eu633 | [X]Trichotillomania | Definite |

| Code | Description | Likelihood |
|---|---|---|
| Eu9.. | [X]Behavioural and emotional disorders with onset usually occurring in childhood and adolescence | Definite |
| Eu900 | [X]Disturbance of activity and attention | Definite |
| Eu91. | [X]Conduct disorders | Definite |
| Eu91z | [X]Conduct disorder, unspecified | Definite |
| Eu92. | [X]Mixed disorders of conduct and emotions | Definite |
| Eu92z | [X]Mixed disorder of conduct and emotions, unspecified | Definite |
| Eu930 | [X]Separation anxiety disorder of childhood | Definite |
| Eu933 | [X]Sibling rivalry disorder | Possible |
| Eu93y | [X]Other childhood emotional disorders | Definite |
| Eu93z | [X]Childhood emotional disorder, unspecified | Definite |
| Eu9y. | [X]Other behavioural and emotional disorders with onset usually occurring in childhood and adolescence | Definite |
| Eu9y7 | [X]Attention deficit disorder | Definite |
| M261E | Acne excoriee des jeunes filles | Possible |
| R0760 | [D]Encopresis NOS | Possible |
| ZV40. | [V]Mental and behavioural problems | Definite |
| ZV403 | [V]Other behavioural problems | Definite |
| ZV405 | [V]Disorder of attention | Definite |
| ZV40y | [V]Other specified mental or behavioural problem | Definite |
| ZV40z | [V]Unspecified mental or behavioural problem | Definite |

##### Appendix A.5.4 Personal and/or Social

Table A8: Read v2 codes contributing to personal and/or social concerns.

| Code | Description | Likelihood |
|---|---|---|
| 1B1J0 | Behavioural, emotional and social difficulties | Definite |
| E2C1. | Nonaggressive unsocial conduct disorder | Definite |
| E2C2. | Socialised conduct disorder | Definite |
| E2D2. | Childhood and adolescent disturbance with sensitivity | Possible |
| E2D20 | Childhood and adolescent disturbance with shyness | Possible |
| Eu932 | [X]Social anxiety disorder of childhood | Definite |
| Eu94. | [X]Disorder of social functioning with onset specific childhood and adolescence | Definite |
| Eu94z | [X]Childhood disorder of social functioning, unspecified | Definite |
| R034C | [D]Social skills development delay | Definite |
| ZVu5B | [X]Inadequate social skills, not elsewhere classified | Definite |

##### Appendix A.5.5 Speech, Language, and/or Communication

Table A9: Read v2 codes contributing to speech, language, and/or communication concerns.

| Code | Description | Likelihood |
|---|---|---|
| 03J6. | Speech therapist | Possible |
| 13Z60 | English as a second language | Possible |
| 13Z68 | Speaks English poorly | Probable |
| 13ZA. | Language difficulty | Definite |
| 13ZA0 | Does not speak English | Definite |
| 13o.. | Communication skills | Possible |
| 13oB. | Difficulty communicating | Definite |

| Code | Description | Likelihood |
|---|---|---|
| 14H2. | H/O: cleft palate | Possible |
| 1B441 | Speech limited | Definite |
| 1B9.. | Speech problem | Definite |
| 1B92. | Has a stammer/stutter | Definite |
| 1B93. | Has difficulty with speech | Definite |
| 1B94. | Speech limited | Definite |
| 1B96. | Speech impairment | Definite |
| 1B9Z. | Speech problem NOS | Definite |
| 1BcX. | Difficulty comprehending speech | Definite |
| 22I30 | Not yet speaking | Definite |
| 2B4.. | O/E - speech defect | Definite |
| 2B48. | O/E - dysarthria | Definite |
| 2B49. | O/E - stammer/stutter | Definite |
| 2B4A. | O/E - speech delay | Definite |
| 2B4Z. | O/E - speech defect NOS | Definite |
| 314.. | Special ENT procedures | Possible |
| 64R3. | Child: speech therapy | Definite |
| 75023 | Unilateral lip adhesion | Possible |
| 7N235 | [SO]Palate | Possible |
| 8E2.. | Speech defect remedial therapy | Definite |
| 8E21. | Speech therapy | Definite |
| 8H7G. | Refer to speech therapist | Probable |
| 8Hl9. | Referral to educational psychologist | Possible |
| 8T0H. | Referral to speech and language therapy service | Probable |
| 9N0Q. | Seen in speech and language clinic | Probable |
| 9N29. | Seen by speech therapist | Probable |
| 9NNj2 | Under care of speech and language therapist | Definite |
| E270. | Stammering or stuttering | Definite |
| E27z2 | Lisping | Definite |
| E2F0z | Specific reading disorder NOS | Definite |
| E2F3. | Speech or language developmental disorder | Definite |
| E2F30 | Developmental aphasia | Definite |
| E2F32 | Articulatory defect due to conductive hearing loss | Definite |
| E2F3z | Speech or language developmental disorder NOS | Definite |
| Eu80. | [X]Specific developmental disorders of speech and language | Definite |
| Eu800 | [X]Specific speech articulation disorder | Definite |
| Eu801 | [X]Expressive language disorder | Definite |
| Eu802 | [X]Receptive language disorder | Definite |
| Eu803 | [X]Acquired aphasia with epilepsy [Landau - Kleffner] | Definite |
| Eu805 | [X]Semantic-pragmatic disorder | Definite |
| Eu80y | [X]Other developmental disorders of speech and language | Definite |
| Eu80z | [X]Developmental disorder of speech and language, unspecified | Definite |
| Eu810 | [X]Specific reading disorder | Definite |
| Eu813 | [X]Mixed disorder of scholastic skills | Possible |
| Eu940 | [X]Elective mutism | Definite |
| Eu951 | [X]Chronic motor or vocal tic disorder | Possible |
| Eu9y5 | [X]Stuttering [stammering] | Definite |
| F143. | Cerebellar ataxia NOS | Possible |
| F23y4 | Ataxic diplegic cerebral palsy | Definite |
| H141. | Tonsil and/or adenoid hypertrophy | Possible |
| H1y3. | Paralysis of vocal cords or larynx | Definite |
| H1y35 | Vocal cord palsy | Definite |
| H1y3z | Laryngoplegia NOS | Definite |
| H1y73 | Stenosis of larynx | Possible |
| P83yX | Congenital malformation of larynx, unspecified | Possible |
| P9... | Cleft palate and lip | Possible |
| P90.. | Cleft palate | Possible |

| Code | Description | Likelihood |
|---|---|---|
| P908. | Incomplete cleft palate NOS | Possible |
| P90z. | Cleft palate NOS | Possible |
| P92.. | Cleft palate with cleft lip | Possible |
| P9z.. | Cleft palate or cleft lip NOS | Possible |
| PA0.. | Tongue tie - ankyloglossia | Probable |
| PAz0. | Unspecified anomalies of mouth and pharynx | Possible |
| PG0C. | Pierre - Robin syndrome | Probable |
| PKy5. | Congenital malformation syndromes affecting facial appearance | Possible |
| Pyu4. | [X]Cleft lip and cleft palate | Possible |
| Pyu41 | [X]Unspecified cleft palate with cleft lip, bilateral | Possible |
| R034A | [D]Communication skills development delay | Definite |
| R043. | [D]Aphasia | Definite |
| R045. | [D]Other speech disturbance | Definite |
| R0452 | [D]Slurred speech | Definite |
| R0454 | [D]Dysfluency | Definite |
| R045z | [D]Other speech disturbance NOS | Definite |
| Ryu6. | [X]Symptoms and signs involving speech and voice | Probable |
| ZV401 | [V]Problems with communication, including speech | Definite |
| ZV459 | [V]Presence of otological and audiological implants | Possible |
| ZV573 | [V]Speech therapy | Definite |

#### Appendix A.5.6 Other Developmental Concerns

Table A10: Read v2 codes contributing to other developmental concerns.

| Code | Description | Likelihood |
|---|---|---|
| 13Z4E | Learning difficulties | Definite |
| 13Z4P | Receiving learning support | Possible |
| 13ZK. | Child with special educational needs | Definite |
| 14g.. | History of developmental disorder | Probable |
| 14g0. | Early childhood developmental disability | Probable |
| 1J9.. | Suspected autism | Probable |
| 22I3. | O/E - delayed milestones | Probable |
| 311B. | Cognitive assessment | Possible |
| 391.. | Feeding ability | Possible |
| 3911. | Needs help with feeding | Possible |
| 3990. | Unable to climb stairs | Possible |
| 8H7T. | Refer to psychologist | Possible |
| 8HT6. | Referral to developmental clinic | Probable |
| 8Hj.. | Referral to education service | Probable |
| 8Hj2. | Referral to psycho-educational group | Possible |
| 8Hkg. | Referral to child development centre | Probable |
| 8Hl9. | Referral to educational psychologist | Possible |
| 8O07. | Provision of special educational needs nursery | Definite |
| 9N0P. | Seen in developmental clinic | Probable |
| 9NNE0 | Under care of educational psychologist | Probable |
| 9NNl. | Under care of autism assessment service | Definite |
| C1zy2 | Cerebral gigantism | Possible |
| C301. | Phenylketonuria | Possible |
| C308. | Disorders of fatty-acid metabolism | Probable |
| C30y8 | Glutaric aciduria Type 1 | Definite |
| C3751 | Mucopolysaccharidosis, type 1 | Definite |

| Code | Description | Likelihood |
|---|---|---|
| C3911 | Di George syndrome | Definite |
| E140. | Infantile autism | Definite |
| E140z | Infantile autism NOS | Definite |
| E273. | Stereotyped repetitive movements | Possible |
| E2F.. | Specific delays in development | Definite |
| E2F0z | Specific reading disorder NOS | Possible |
| E2F2. | Other specific learning difficulty | Definite |
| E2F30 | Developmental aphasia | Definite |
| E2F5. | Mixed development disorder | Definite |
| E2Fy. | Other development delays | Definite |
| E2Fz. | Developmental disorder NOS | Definite |
| Eu057 | [X]Mild cognitive disorder | Definite |
| Eu8.. | [X]Disorders of psychological development | Definite |
| Eu80y | [X]Other developmental disorders of speech and language | Definite |
| Eu80z | [X]Developmental disorder of speech and language, unspecified | Definite |
| Eu81z | [X]Developmental disorder of scholastic skills, unspecified | Definite |
| Eu83. | [X]Mixed specific developmental disorders | Definite |
| Eu84. | [X]Pervasive developmental disorders | Definite |
| Eu840 | [X]Childhood autism | Definite |
| Eu842 | [X]Rett's syndrome | Definite |
| Eu845 | [X]Asperger's syndrome | Definite |
| Eu84z | [X]Pervasive developmental disorder, unspecified | Definite |
| Eu85. | [X]Global developmental delay | Definite |
| Eu86. | [X]Neurodevelopmental delay | Definite |
| K0802 | Renal infantilism | Definite |
| P223. | Agyria | Definite |
| P226. | Microgyria | Definite |
| P2283 | Aicardi syndrome | Definite |
| P22y3 | Partial absence of septum pellucidum | Possible |
| P246. | Septo-optic dysplasia | Definite |
| PJ1.. | Patau's syndrome - trisomy 13 | Definite |
| PJ2.. | Edward's syndrome - trisomy 18 | Definite |
| PJ333 | Smith-Magenis syndrome | Definite |
| PJ9.. | Mowat-Wilson syndrome | Definite |
| PJy2. | XXX syndrome | Definite |
| PJy3. | XXY syndrome | Definite |
| PJyy2 | Fragile X chromosome | Definite |
| PJyy3 | Karyotype 47,XYY | Possible |
| PJyy4 | Fragile X syndrome | Definite |
| PJz0. | Mosaicism NOS | Definite |
| PK80. | Fetal alcohol syndrome | Definite |
| PK84. | Fetal valproate syndrome | Definite |
| PKy1. | Laurence-Moon-Biedl syndrome | Definite |
| PKy4. | William syndrome | Definite |
| PKy5D | Kabuki make-up syndrome | Definite |
| PKy5K | Cohen syndrome | Probable |
| PKy62 | Russell - Silver syndrome | Probable |
| PKy73 | Rubenstein - Tayi syndrome | Definite |
| PKy93 | Prader - Willi syndrome | Definite |
| PKyD. | Nicolaides-Baraitser syndrome | Definite |
| PKyz6 | Congenital hemihypertrophy | Probable |
| PKyz7 | Angelman's syndrome | Definite |
| R034. | [D]Physiological development failure | Probable |
| R0340 | [D]Delayed milestone | Probable |
| R0344 | [D]Physical retardation | Definite |
| R034B | [D]Fine motor skills development delay | Definite |
| R034C | [D]Social skills development delay | Probable |

| Code | Description | Likelihood |
|---|---|---|
| R034E | [D]Developmental delay | Definite |
| ZV400 | [V]Problems with learning | Definite |
| ZV57E | [V] Admission for toilet training | Possible |
| ZV793 | [V]Screening for early childhood developmental handicap | Possible |

### Appendix B Psychosocial Health Outcome Prevalence

#### Appendix B.1 Children in Care Compared With Children Not in Care

Table A11: Sensitivity analysis: Prevalence and relative risks for psychosocial concerns among children in care and children not in care

| | In care N = 7887 | | | Not in care N = 445 547 | | | Relative risk In care v. Not in care | |
|---|---|---|---|---|---|---|---|---|
| Outcome | N | % | 95% CI | N | % | 95% CI | RR | 95% CI |
| Any psychosocial | 2541 | 32.2 | 31.2–33.3 | 95232 | 21.4 | 21.3–21.5 | 1.51 | 1.46–1.56 |
| EBA | 1345 | 17.1 | 16.2–17.9 | 35207 | 7.9 | 7.8–8.0 | 2.16 | 2.05–2.27 |
| Personal and/or social | 1134 | 14.4 | 13.6–15.2 | 41093 | 9.2 | 9.1–9.3 | 1.56 | 1.48–1.65 |
| SLC | 1861 | 23.6 | 22.7–24.5 | 73843 | 16.6 | 16.5–16.7 | 1.42 | 1.37–1.48 |
| Other developmental | 115 | 1.5 | 1.2–1.7 | 3393 | 0.8 | 0.7–0.8 | 1.91 | 1.59–2.30 |

The "all potential cases" definition used in sensitivity analyses includes indicators rated *definite*, *probable*, or *possible*. Values are prevalence estimates with 95% confidence intervals and unadjusted relative risks comparing children in care with children not in care. "Any psychosocial" indicates a concern in any outcome listed. RR = relative risk. EBA = emotional, behavioral, and/or attentional. SLC = speech, language, and/or communication.

#### Appendix B.2 Children With Unknown Care Status Compared With Children Not in Care

Table A12: Primary analysis: Prevalence and relative risks for psychosocial concerns among children with unknown care status and children not in care

| | Unknown care status N = 23 730 | | | Not in care N = 445 547 | | | Relative risk Unknown v. Not in care | |
|---|---|---|---|---|---|---|---|---|
| Outcome | N | % | 95% CI | N | % | 95% CI | RR | 95% CI |
| Any psychosocial | 3407 | 14.4 | 13.9–14.8 | 77836 | 17.5 | 17.4–17.6 | 0.82 | 0.80–0.85 |
| EBA | 1082 | 4.6 | 4.3–4.8 | 29208 | 6.6 | 6.5–6.6 | 0.70 | 0.66–0.74 |
| Personal and/or social | 863 | 3.6 | 3.4–3.9 | 23416 | 5.3 | 5.2–5.3 | 0.69 | 0.65–0.74 |
| SLC | 2928 | 12.3 | 11.9–12.8 | 63989 | 14.4 | 14.3–14.5 | 0.86 | 0.83–0.89 |
| Other developmental | 116 | 0.5 | 0.4–0.6 | 3363 | 0.8 | 0.7–0.8 | 0.65 | 0.54–0.78 |

The “likely cases” definition used in primary analyses includes indicators rated *definite* or *probable*. Values are prevalence estimates with 95% confidence intervals and unadjusted relative risks comparing children with unknown care status with children not in care. “Any psychosocial” indicates a concern in any outcome listed. RR = relative risk. EBA = emotional, behavioral, and/or attentional. SLC = speech, language, and/or communication.

Table A13: Sensitivity analysis: Prevalence and relative risks for psychosocial concerns among children with unknown care status and children not in care

| | Unknown care status N = 23 730 | | | Not in care N = 445 547 | | | Relative risk Unknown v. Not in care | |
|---|---|---|---|---|---|---|---|---|
| Outcome | N | % | 95% CI | N | % | 95% CI | RR | 95% CI |
| Any psychosocial | 4122 | 17.4 | 16.9–17.9 | 95232 | 21.4 | 21.3–21.5 | 0.81 | 0.79–0.84 |
| EBA | 1266 | 5.3 | 5.1–5.6 | 35207 | 7.9 | 7.8–8.0 | 0.68 | 0.64–0.71 |
| Personal and/or social | 1402 | 5.9 | 5.6–6.2 | 41093 | 9.2 | 9.1–9.3 | 0.64 | 0.61–0.67 |
| SLC | 3425 | 14.4 | 14.0–14.9 | 73843 | 16.6 | 16.5–16.7 | 0.87 | 0.84–0.90 |
| Other developmental | 117 | 0.5 | 0.4–0.6 | 3393 | 0.8 | 0.7–0.8 | 0.65 | 0.54–0.78 |

The “all potential cases” definition used in sensitivity analyses includes indicators rated *definite*, *probable*, or *possible*. Values are prevalence estimates with 95% confidence intervals and unadjusted relative risks comparing children with unknown care status with children not in care. “Any psychosocial” indicates a concern in any outcome listed. RR = relative risk. EBA = emotional, behavioral, and/or attentional. SLC = speech, language, and/or communication.

#### Appendix B.3 Children in Care Compared With Children With Unknown Care Status

Table A14: Primary analysis: Prevalence and relative risks for psychosocial concerns among children in care and children with unknown care status

| | In care N = 7887 | | | Unknown care status N = 23 730 | | | Relative risk In care v. Unknown | |
|---|---|---|---|---|---|---|---|---|
| Outcome | N | % | 95% CI | N | % | 95% CI | RR | 95% CI |
| Any psychosocial | 2290 | 29.0 | 28.0–30.0 | 3407 | 14.4 | 13.9–14.8 | 2.02 | 1.93–2.12 |
| EBA | 1224 | 15.5 | 14.7–16.3 | 1082 | 4.6 | 4.3–4.8 | 3.40 | 3.15–3.68 |
| Personal and/or social | 834 | 10.6 | 9.9–11.3 | 863 | 3.6 | 3.4–3.9 | 2.91 | 2.65–3.19 |
| SLC | 1706 | 21.6 | 20.7–22.6 | 2928 | 12.3 | 11.9–12.8 | 1.75 | 1.66–1.85 |
| Other developmental | 114 | 1.4 | 1.2–1.7 | 116 | 0.5 | 0.4–0.6 | 2.96 | 2.29–3.82 |

The “likely cases” definition used in primary analyses includes indicators rated *definite* or *probable*. Values are prevalence estimates with 95% confidence intervals and unadjusted relative risks comparing children in care with children with unknown care status. “Any psychosocial” indicates a concern in any outcome listed. RR = relative risk. EBA = emotional, behavioral, and/or attentional. SLC = speech, language, and/or communication.

Table A15: Sensitivity analysis: Prevalence and relative risks for psychosocial concerns among children in care and children with unknown care status

| | In care N = 7887 | | | Unknown care status N = 23 730 | | | Relative risk In care v. Unknown | |
|---|---|---|---|---|---|---|---|---|
| Outcome | N | % | 95% CI | N | % | 95% CI | RR | 95% CI |
| Any psychosocial | 2541 | 32.2 | 31.2–33.3 | 4122 | 17.4 | 16.9–17.9 | 1.85 | 1.78–1.94 |
| EBA | 1345 | 17.1 | 16.2–17.9 | 1266 | 5.3 | 5.1–5.6 | 3.20 | 2.97–3.44 |
| Personal and/or social | 1134 | 14.4 | 13.6–15.2 | 1402 | 5.9 | 5.6–6.2 | 2.43 | 2.26–2.62 |
| SLC | 1861 | 23.6 | 22.7–24.5 | 3425 | 14.4 | 14.0–14.9 | 1.63 | 1.55–1.72 |
| Other developmental | 115 | 1.5 | 1.2–1.7 | 117 | 0.5 | 0.4–0.6 | 2.96 | 2.29–3.82 |

The “all potential cases” definition used in sensitivity analyses includes indicators rated *definite*, *probable*, or *possible*. Values are prevalence estimates with 95% confidence intervals and unadjusted relative risks comparing children in care with children with unknown care status. “Any psychosocial” indicates a concern in any outcome listed. RR = relative risk. EBA = emotional, behavioral, and/or attentional. SLC = speech, language, and/or communication.

#### Appendix B.4 Prevalence Stratified by Care Status Across Deprivation Groups

Figure A1: Sensitivity analysis: Deprivation gradient of prevalence of any psychosocial health concern by care status and relative risk for children in care compared to children not in care

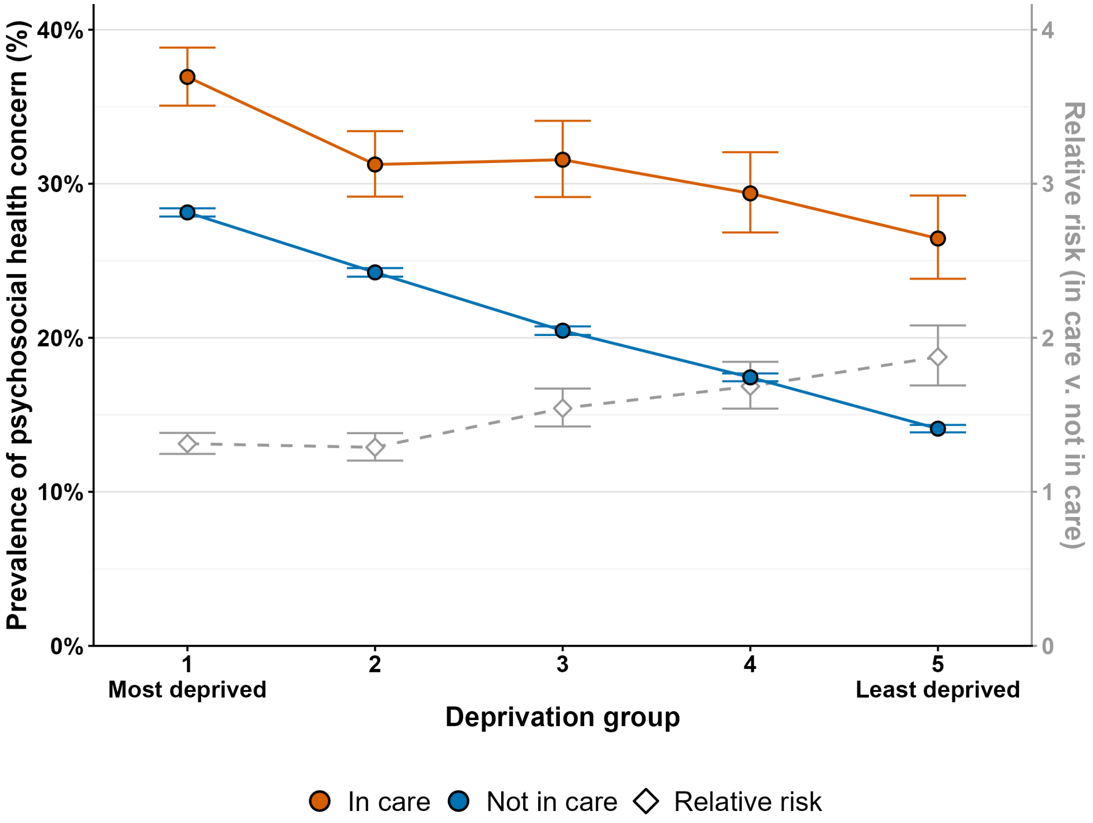


The "all potential cases" definition used in sensitivity analyses includes indicators rated *definite*, *probable*, or *possible*. Circular points show prevalence estimates and 95% confidence intervals. Diamond points and the dashed line show unadjusted relative risks and 95% confidence intervals comparing children in care with children not in care within each deprivation group, using the right-hand vertical axis.

Figure A2: Primary analysis: Deprivation gradient of prevalence of emotional, behavioral, and/or attentional concern by care status, and relative risk for children in care compared to children not in care

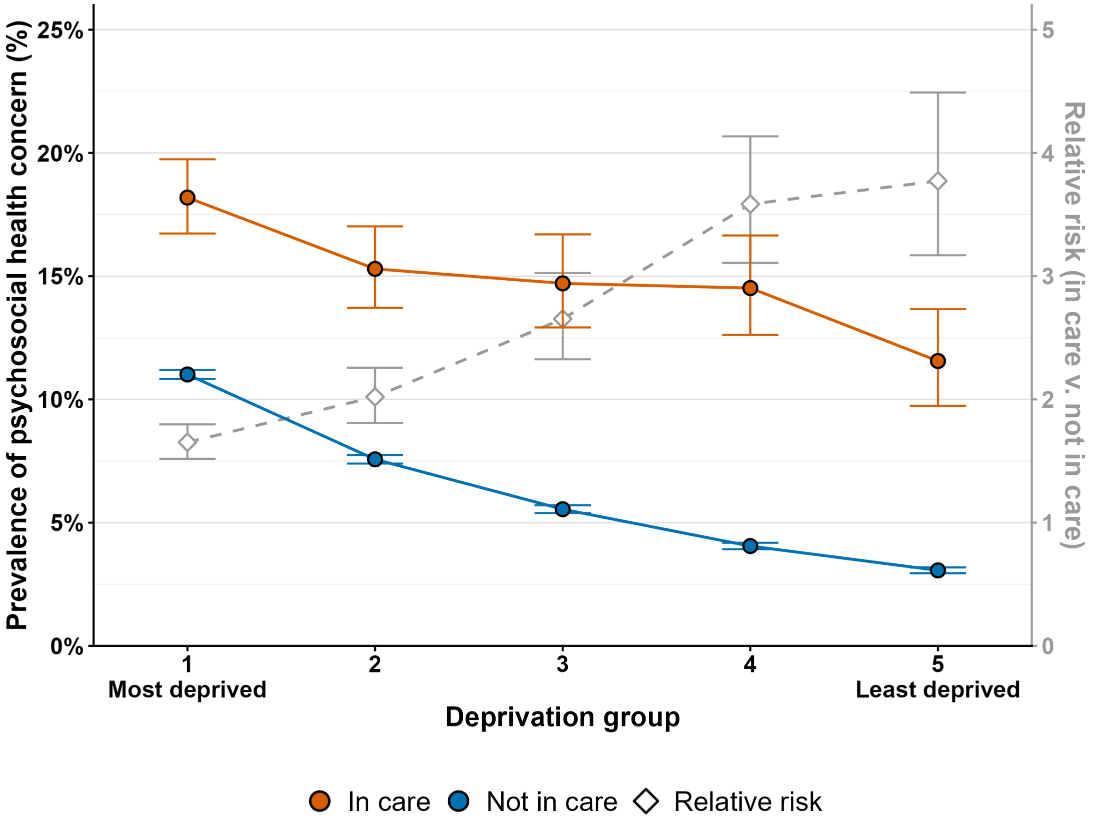


The “likely cases” definition used in primary analyses includes indicators rated *definite* or *probable*. Circular points show prevalence estimates and 95% confidence intervals. Diamond points and the dashed line show unadjusted relative risks and 95% confidence intervals comparing children in care with children not in care within each deprivation group, using the right-hand vertical axis.

Figure A3: Primary analysis: Deprivation gradient of prevalence of personal and/or social concern by care status and relative risk for children in care compared to children not in care

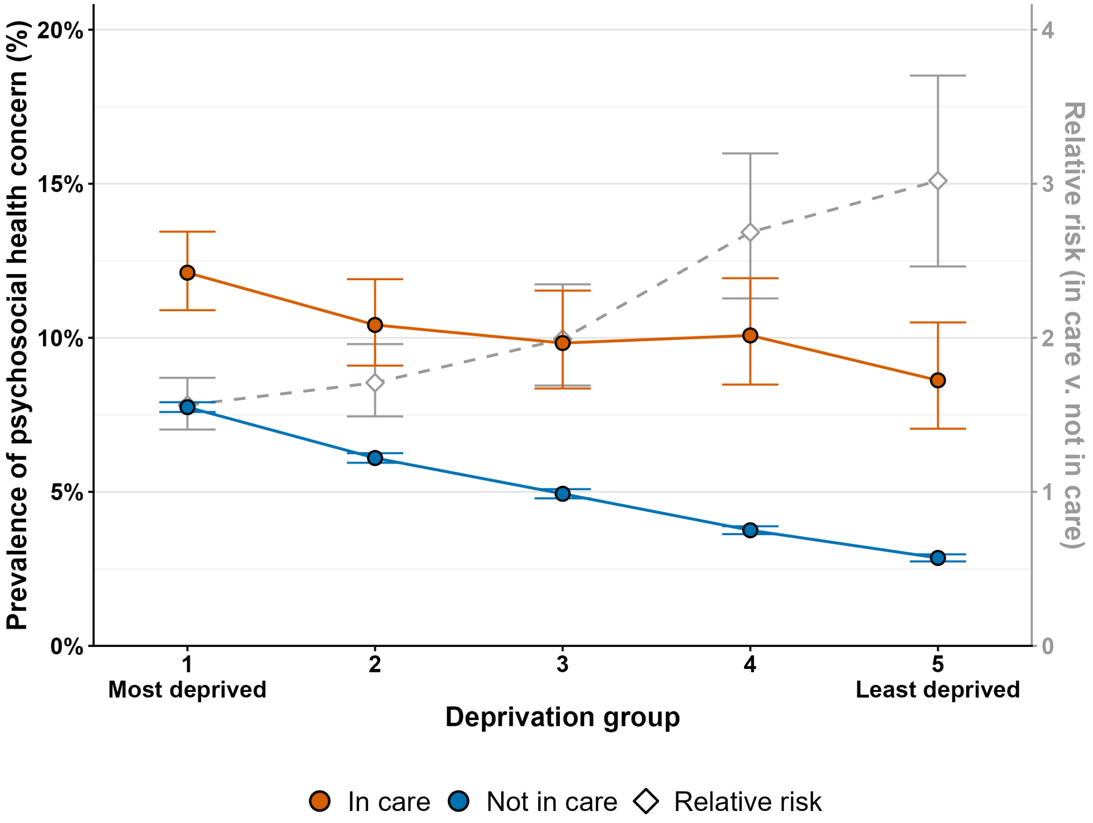


The "likely cases" definition used in primary analyses includes indicators rated *definite* or *probable*. Circular points show prevalence estimates and 95% confidence intervals. Diamond points and the dashed line show unadjusted relative risks and 95% confidence intervals comparing children in care with children not in care within each deprivation group, using the right-hand vertical axis.

Figure A4: Primary analysis: Deprivation gradient of prevalence of speech, language, and/or communication concern by care status and relative risk for children in care compared to children not in care

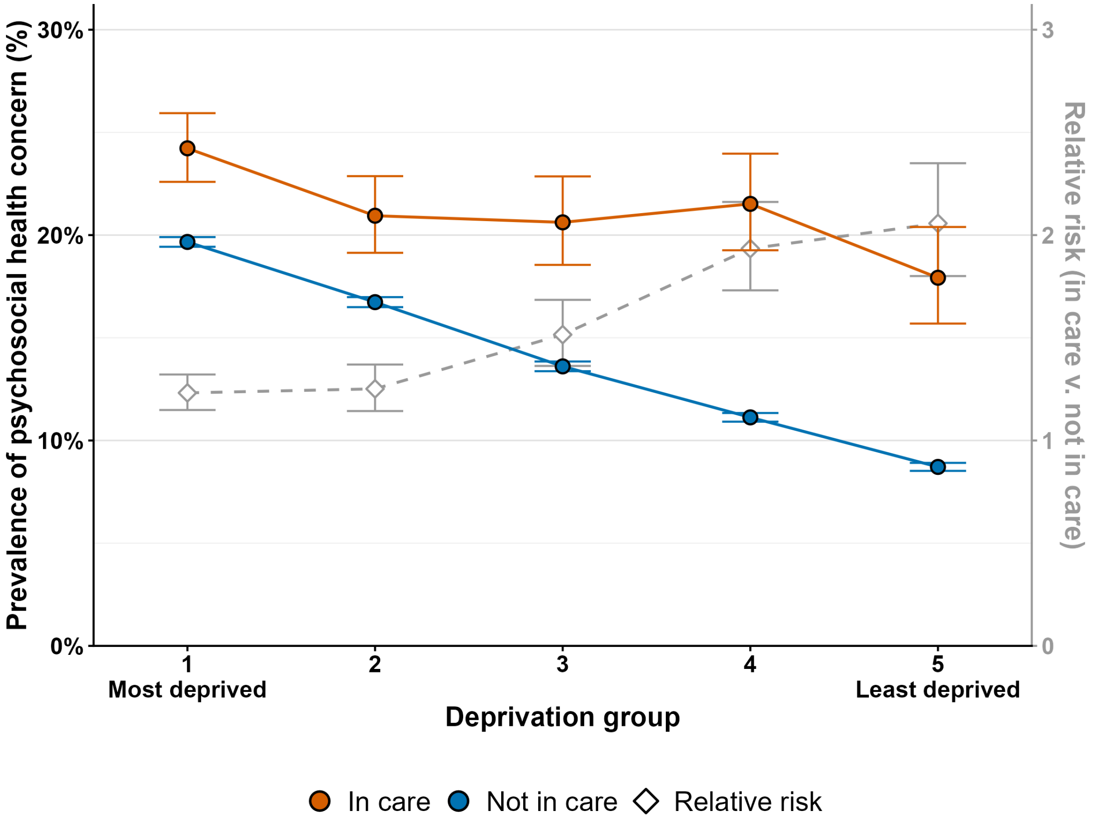


The “likely cases” definition used in primary analyses includes indicators rated *definite* or *probable*. Circular points show prevalence estimates and 95% confidence intervals. Diamond points and the dashed line show unadjusted relative risks and 95% confidence intervals comparing children in care with children not in care within each deprivation group, using the right-hand vertical axis.

Figure A5: Primary analysis: Prevalence of other developmental concern by deprivation group and care status, with relative risks comparing children in care with children not in care

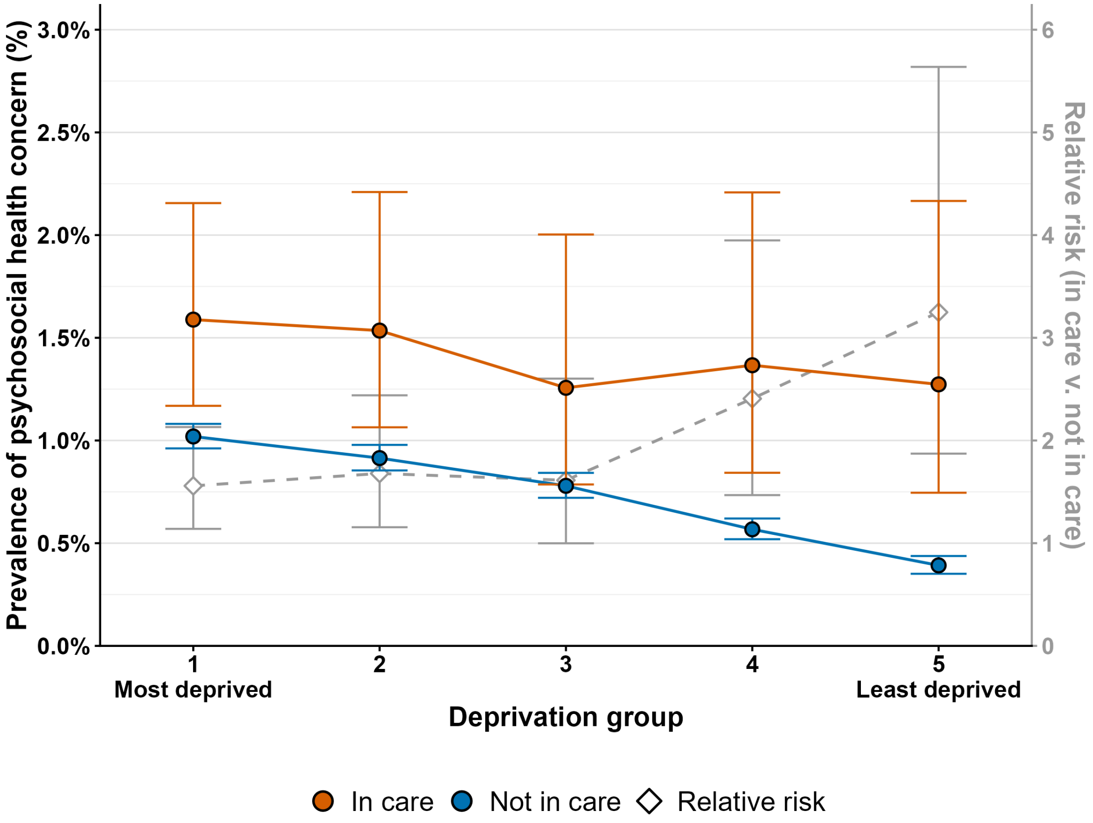


The “likely cases” definition used in primary analyses includes indicators rated *definite* or *probable*. Circular points show prevalence estimates and 95% confidence intervals. Diamond points and the dashed line show unadjusted relative risks and 95% confidence intervals comparing children in care with children not in care within each deprivation group, using the right-hand vertical axis.

### Appendix C Regression Models and Contrasts

#### Appendix C.1 Supplementary Regression Tables

Model coefficients and contrast estimates are reported below, with one table per outcome. Tables A16–A20 show the development of model coefficients with increasing model complexity. Each table shows odds ratios and 95% confidence intervals from three model specifications: univariable models, an adjusted main-effects model, and an adjusted model including a care status by deprivation group interaction. Raw interaction coefficients are not reported because these are model parameters rather than the subgroup contrasts of substantive interest. The relevant model-derived contrasts comparing children in care with children not in care within each deprivation group are reported separately in Tables A21–A25. This is because these adjusted odds ratios are calculated using combinations of linear coefficients. Similarly, the adjusted odds ratios between deprivation groups within care-status groups are reported in Tables A26–A30.

Table A16: Logistic regression estimates and model comparison for any psychosocial concern

| **Variable** | **(1) Univariable** | **(2) Adjusted** | **(3) Adjusted + interaction** |
|---|---|---|---|
| | **Care status** | | |
| In care | 1.93 (1.84–2.03) | 1.86 (1.77–1.96) | |
| Not in care (ref) | 1.00 | 1.00 | *See Table A21* |
| Unknown | 0.79 (0.76–0.82) | 0.84 (0.81–0.87) | |
| | **Deprivation group** | | |
| Dep. group 1 - Most dep. | 2.79 (2.72–2.87) | 2.81 (2.74–2.89) | |
| Dep. group 2 | 2.13 (2.08–2.19) | 2.16 (2.11–2.22) | |
| Dep. group 3 | 1.64 (1.59–1.69) | 1.66 (1.61–1.71) | *See Table A26* |
| Dep. group 4 | 1.30 (1.26–1.34) | 1.32 (1.28–1.36) | |
| Dep. group 5 - Least dep. (ref) | 1.00 | 1.00 | |
| | **Other covariates** | | |
| Age, per week | 1.007 (1.006–1.008) | 1.001 (1.000–1.002) | 1.001 (1.000–1.002) |
| Sex | | | |
| Female (ref) | 1.00 | 1.00 | 1.00 |
| Male | 2.21 (2.17–2.24) | 2.25 (2.21–2.28) | 2.25 (2.21–2.28) |
| Ethnicity | | | |
| White (ref) | 1.00 | 1.00 | 1.00 |
| Other ethnic group | 1.31 (1.28–1.34) | 1.27 (1.24–1.31) | 1.27 (1.24–1.31) |
| | **Model comparison** | | |
| Likelihood-ratio test | – | Reference model | $\chi^2_8 = 86.96, p < 0.001$ |

Values are odds ratios and 95% confidence intervals. Model (1) are univariable models; model (2) adjusts for care status, deprivation group, age at review, sex, and ethnicity; model (3) additionally includes a care status by deprivation group interaction. For model (3), care-status contrasts are reported in Table A21 and deprivation contrasts in Table A26. The likelihood-ratio test compares models (2) and (3).

Table A17: Logistic regression estimates and model comparison for emotional, behavioral, and/or attentional concern

| Variable | (1) Univariable | (2) Adjusted | (3) Adjusted + interaction |
|---|---|---|---|
| **Care status** | | | |
| In care | 2.62 (2.46–2.79) | 2.49 (2.34–2.66) | |
| Not in care (ref) | 1.00 | 1.00 | *See Table A22* |
| Unknown | 0.68 (0.64–0.72) | 0.75 (0.71–0.80) | |
| **Deprivation group** | | | |
| Dep. group 1 - Most dep. | 3.86 (3.70–4.03) | 3.72 (3.56–3.88) | |
| Dep. group 2 | 2.56 (2.45–2.68) | 2.56 (2.44–2.67) | |
| Dep. group 3 | 1.85 (1.76–1.94) | 1.86 (1.78–1.96) | *See Table A27* |
| Dep. group 4 | 1.35 (1.29–1.42) | 1.37 (1.30–1.44) | |
| Dep. group 5 - Least dep. (ref) | 1.00 | 1.00 | |
| **Other covariates** | | | |
| Age, per week | 1.023 (1.022–1.025) | 1.016 (1.014–1.018) | 1.016 (1.014–1.018) |
| Sex | | | |
| Female (ref) | 1.00 | 1.00 | 1.00 |
| Male | 2.16 (2.11–2.22) | 2.19 (2.14–2.25) | 2.19 (2.14–2.25) |
| Ethnicity | | | |
| White (ref) | 1.00 | 1.00 | 1.00 |
| Other ethnic group | 1.26 (1.22–1.31) | 1.15 (1.11–1.20) | 1.15 (1.11–1.20) |
| **Model comparison** | | | |
| Likelihood-ratio test | – | Reference model | $\chi^2_8 = 115.78, p < 0.001$ |

Values are odds ratios and 95% confidence intervals. Model (1) are univariable models; model (2) adjusts for care status, deprivation group, age at review, sex, and ethnicity; model (3) additionally includes a care status by deprivation group interaction. For model (3), care-status contrasts are reported in Table A22 and deprivation contrasts in Table A27. The likelihood-ratio test compares models (2) and (3).

Table A18: Logistic regression estimates and model comparison for personal and/or social concern

| **Variable** | **(1) Univariable** | **(2) Adjusted** | **(3) Adjusted + interaction** |
|---|---|---|---|
| **Care status** | | | |
| In care | 2.13 (1.98–2.29) | 2.04 (1.90–2.20) | |
| Not in care (ref) | 1.00 | 1.00 | *See Table A23* |
| Unknown | 0.68 (0.63–0.73) | 0.72 (0.67–0.77) | |
| **Deprivation group** | | | |
| Dep. group 1 - Most dep. | 2.86 (2.73–2.99) | 2.84 (2.71–2.97) | |
| Dep. group 2 | 2.20 (2.09–2.30) | 2.21 (2.11–2.32) | |
| Dep. group 3 | 1.75 (1.67–1.85) | 1.78 (1.69–1.87) | *See Table A28* |
| Dep. group 4 | 1.33 (1.26–1.40) | 1.35 (1.28–1.42) | |
| Dep. group 5 - Least dep. (ref) | 1.00 | 1.00 | |
| **Other covariates** | | | |
| Age, per week | 1.005 (1.003–1.006) | 0.997 (0.996–0.999) | 0.997 (0.996–0.999) |
| Sex | | | |
| Female (ref) | 1.00 | 1.00 | 1.00 |
| Male | 2.66 (2.58–2.74) | 2.68 (2.61–2.76) | 2.68 (2.61–2.76) |
| Ethnicity | | | |
| White (ref) | 1.00 | 1.00 | 1.00 |
| Other ethnic group | 1.48 (1.43–1.54) | 1.46 (1.40–1.52) | 1.46 (1.40–1.52) |
| **Model comparison** | | | |
| Likelihood-ratio test | – | Reference model | $\chi^2_8 = 43.54$, $p < 0.001$ |

Values are odds ratios and 95% confidence intervals. Model (1) are univariable models; model (2) adjusts for care status, deprivation group, age at review, sex, and ethnicity; model (3) additionally includes a care status by deprivation group interaction. For model (3), care-status contrasts are reported in Table A23 and deprivation contrasts in Table A28. The likelihood-ratio test compares models (2) and (3).

Table A19: Logistic regression estimates and model comparison for speech, language, and/or communication concern

| Variable | (1) Univariable | (2) Adjusted | (3) Adjusted + interaction |
|---|---|---|---|
| **Care status** | | | |
| In care | 1.65 (1.56–1.74) | 1.57 (1.49–1.66) | |
| Not in care (ref) | 1.00 | 1.00 | *See Table A24* |
| Unknown | 0.84 (0.81–0.87) | 0.87 (0.84–0.91) | |
| **Deprivation group** | | | |
| Dep. group 1 - Most dep. | 2.57 (2.50–2.64) | 2.62 (2.55–2.70) | |
| Dep. group 2 | 2.10 (2.04–2.16) | 2.14 (2.07–2.20) | |
| Dep. group 3 | 1.64 (1.59–1.69) | 1.66 (1.61–1.71) | *See Table A29* |
| Dep. group 4 | 1.32 (1.28–1.36) | 1.34 (1.29–1.38) | |
| Dep. group 5 - Least dep. (ref) | 1.00 | 1.00 | |
| **Other covariates** | | | |
| Age, per week | 0.999 (0.998–1.000) | 0.993 (0.992–0.994) | 0.993 (0.992–0.994) |
| Sex | | | |
| Female (ref) | 1.00 | 1.00 | 1.00 |
| Male | 2.28 (2.24–2.32) | 2.31 (2.27–2.35) | 2.31 (2.27–2.35) |
| Ethnicity | | | |
| White (ref) | 1.00 | 1.00 | 1.00 |
| Other ethnic group | 1.31 (1.28–1.35) | 1.31 (1.28–1.35) | 1.31 (1.28–1.35) |
| **Model comparison** | | | |
| Likelihood-ratio test | – | Reference model | $\chi^2_8 = 83.37, p < 0.001$ |

Values are odds ratios and 95% confidence intervals. Model (1) are univariable models; model (2) adjusts for care status, deprivation group, age at review, sex, and ethnicity; model (3) additionally includes a care status by deprivation group interaction. For model (3), care-status contrasts are reported in Table A24 and deprivation contrasts in Table A29. The likelihood-ratio test compares models (2) and (3).

Table A20: Logistic regression estimates and model comparison for other developmental concern

| Variable | (1) Univariable | (2) Adjusted | (3) Adjusted + interaction |
|---|---|---|---|
| **Care status** | | | |
| In care | 1.93 (1.60–2.33) | 1.83 (1.52–2.21) | |
| Not in care (ref) | 1.00 | 1.00 | *See Table A25* |
| Unknown | 0.65 (0.54–0.78) | 0.67 (0.55–0.80) | |
| **Deprivation group** | | | |
| Dep. group 1 - Most dep. | 2.53 (2.25–2.86) | 2.52 (2.24–2.85) | |
| Dep. group 2 | 2.25 (1.99–2.55) | 2.26 (1.99–2.56) | |
| Dep. group 3 | 1.91 (1.68–2.17) | 1.92 (1.69–2.19) | *See Table A30* |
| Dep. group 4 | 1.41 (1.23–1.62) | 1.43 (1.25–1.64) | |
| Dep. group 5 - Least dep. (ref) | 1.00 | 1.00 | |
| **Other covariates** | | | |
| Age, per week | 0.997 (0.992–1.001) | 0.991 (0.986–0.996) | 0.991 (0.986–0.996) |
| Sex | | | |
| Female (ref) | 1.00 | 1.00 | 1.00 |
| Male | 2.35 (2.18–2.52) | 2.35 (2.19–2.53) | 2.35 (2.19–2.53) |
| Ethnicity | | | |
| White (ref) | 1.00 | 1.00 | 1.00 |
| Other ethnic group | 1.40 (1.27–1.54) | 1.41 (1.28–1.56) | 1.41 (1.28–1.56) |
| **Model comparison** | | | |
| Likelihood-ratio test | – | Reference model | $\chi^2_8 = 10.85$, $p = 0.210$ |

Values are odds ratios and 95% confidence intervals. Model (1) are univariable models; model (2) adjusts for care status, deprivation group, age at review, sex, and ethnicity; model (3) additionally includes a care status by deprivation group interaction. For model (3), care-status contrasts are reported in Table A25 and deprivation contrasts in Table A30. The likelihood-ratio test compares models (2) and (3).

#### Appendix C.2 Care-Status Contrasts Within Deprivation Groups

The following tables report adjusted odds ratios arising from model (3). They compare children in care with children not in care across deprivation groups.

Table A21: Adjusted odds ratios comparing children in care with children not in care within deprivation groups for any psychosocial concern

| **Deprivation group** | **aOR (95% CI)** |
|---|---|
| 1 - Most deprived | 1.58 (1.45–1.72) |
| 2 | 1.63 (1.47–1.81) |
| 3 | 1.99 (1.76–2.25) |
| 4 | 2.61 (2.29–2.98) |
| 5 - Least deprived | 2.61 (2.25–3.03) |

Estimates are derived from model (3), which adjusted for age at review, sex, and ethnicity and included a care status by deprivation group interaction. aOR = adjusted odds ratio.

Table A22: Adjusted odds ratios comparing children in care with children not in care within deprivation groups for emotional, behavioral, and/or attentional concern

| **Deprivation group** | **aOR (95% CI)** |
|---|---|
| 1 - Most deprived | 1.85 (1.67–2.05) |
| 2 | 2.27 (1.99–2.59) |
| 3 | 3.05 (2.61–3.56) |
| 4 | 4.20 (3.55–4.96) |
| 5 - Least deprived | 4.29 (3.52–5.23) |

Estimates are derived from model (3), which adjusted for age at review, sex, and ethnicity and included a care status by deprivation group interaction. aOR = adjusted odds ratio.

Table A23: Adjusted odds ratios comparing children in care with children not in care within deprivation groups for personal and/or social concern

| **Deprivation group** | **aOR (95% CI)** |
|---|---|
| 1 - Most deprived | 1.69 (1.49–1.91) |
| 2 | 1.84 (1.58–2.15) |
| 3 | 2.16 (1.80–2.59) |
| 4 | 2.96 (2.43–3.60) |
| 5 - Least deprived | 3.31 (2.65–4.15) |

Estimates are derived from model (3), which adjusted for age at review, sex, and ethnicity and included a care status by deprivation group interaction. aOR = adjusted odds ratio.

Table A24: Adjusted odds ratios comparing children in care with children not in care within deprivation groups for speech, language, and/or communication concern

| Deprivation group | aOR (95% CI) |
|---|---|
| 1 - Most deprived | 1.33 (1.21–1.46) |
| 2 | 1.34 (1.19–1.50) |
| 3 | 1.68 (1.47–1.93) |
| 4 | 2.24 (1.94–2.59) |
| 5 - Least deprived | 2.34 (1.99–2.76) |

Estimates are derived from model (3), which adjusted for age at review, sex, and ethnicity and included a care status by deprivation group interaction. aOR = adjusted odds ratio.

Table A25: Adjusted odds ratios comparing children in care with children not in care within deprivation groups for other developmental concern

| Deprivation group | aOR (95% CI) |
|---|---|
| 1 - Most deprived | 1.59 (1.15–2.18) |
| 2 | 1.71 (1.17–2.50) |
| 3 | 1.64 (1.01–2.66) |
| 4 | 2.44 (1.48–4.04) |
| 5 - Least deprived | 3.32 (1.90–5.80) |

Estimates are derived from model (3), which adjusted for age at review, sex, and ethnicity and included a care status by deprivation group interaction. aOR = adjusted odds ratio.

#### Appendix C.3 Deprivation Contrasts Within Care-Status Groups

The following tables report adjusted odds ratios arising from model (3). They compare deprivation groups separately for children in care and children not in care.

Table A26: Adjusted odds ratios comparing deprivation groups within care status groups for any psychosocial concern

| Deprivation group | In care, aOR (95% CI) | Not in care, aOR (95% CI) |
|---|---|---|
| 1 - Most deprived | 1.71 (1.44–2.03) | 2.82 (2.75–2.90) |
| 2 | 1.36 (1.13–1.62) | 2.17 (2.11–2.23) |
| 3 | 1.27 (1.05–1.54) | 1.67 (1.62–1.72) |
| 4 | 1.30 (1.07–1.59) | 1.30 (1.26–1.34) |
| 5 - Least deprived (ref) | 1.00 | 1.00 |

Rows for deprivation groups 1 to 4 are adjusted odds ratios comparing that group with deprivation group 5 separately among children in care and children not in care. Estimates are derived from model (3), which adjusted for age at review, sex, and ethnicity and included a care status by deprivation group interaction. aOR = adjusted odds ratio.

Table A27: Adjusted odds ratios comparing deprivation groups within care status groups for emotional, behavioral, and/or attentional concern

| Deprivation group | In care, aOR (95% CI) | Not in care, aOR (95% CI) |
|---|---|---|
| 1 - Most deprived | 1.65 (1.32–2.05) | 3.83 (3.66–4.00) |
| 2 | 1.38 (1.09–1.74) | 2.61 (2.49–2.73) |
| 3 | 1.33 (1.04–1.70) | 1.88 (1.79–1.98) |
| 4 | 1.32 (1.02–1.70) | 1.35 (1.28–1.42) |
| 5 - Least deprived (ref) | 1.00 | 1.00 |

Rows for deprivation groups 1 to 4 are adjusted odds ratios comparing that group with deprivation group 5 separately among children in care and children not in care. Estimates are derived from model (3), which adjusted for age at review, sex, and ethnicity and included a care status by deprivation group interaction. aOR = adjusted odds ratio.

Table A28: Adjusted odds ratios comparing deprivation groups within care status groups for personal and/or social concern

| Deprivation group | In care, aOR (95% CI) | Not in care, aOR (95% CI) |
|---|---|---|
| 1 - Most deprived | 1.46 (1.14–1.88) | 2.87 (2.74–3.01) |
| 2 | 1.24 (0.95–1.62) | 2.24 (2.13–2.35) |
| 3 | 1.16 (0.88–1.55) | 1.79 (1.70–1.89) |
| 4 | 1.20 (0.89–1.61) | 1.34 (1.27–1.42) |
| 5 - Least deprived (ref) | 1.00 | 1.00 |

Rows for deprivation groups 1 to 4 are adjusted odds ratios comparing that group with deprivation group 5 separately among children in care and children not in care. Estimates are derived from model (3), which adjusted for age at review, sex, and ethnicity and included a care status by deprivation group interaction. aOR = adjusted odds ratio.

Table A29: Adjusted odds ratios comparing deprivation groups within care status groups for speech, language, and/or communication concern

| Deprivation group | In care, aOR (95% CI) | Not in care, aOR (95% CI) |
|---|---|---|
| 1 - Most deprived | 1.49 (1.24–1.80) | 2.63 (2.56–2.71) |
| 2 | 1.23 (1.01–1.50) | 2.14 (2.08–2.21) |
| 3 | 1.20 (0.97–1.48) | 1.67 (1.62–1.72) |
| 4 | 1.27 (1.02–1.57) | 1.32 (1.28–1.37) |
| 5 - Least deprived (ref) | 1.00 | 1.00 |

Rows for deprivation groups 1 to 4 are adjusted odds ratios comparing that group with deprivation group 5 separately among children in care and children not in care. Estimates are derived from model (3), which adjusted for age at review, sex, and ethnicity and included a care status by deprivation group interaction. aOR = adjusted odds ratio.

Table A30: Adjusted odds ratios comparing deprivation groups within care status groups for other developmental concern

| Deprivation group | In care, aOR (95% CI) | Not in care, aOR (95% CI) |
|---|---|---|
| 1 - Most deprived | 1.26 (0.67–2.38) | 2.64 (2.33–3.00) |
| 2 | 1.22 (0.63–2.36) | 2.36 (2.07–2.69) |
| 3 | 0.99 (0.48–2.05) | 2.01 (1.75–2.30) |
| 4 | 1.08 (0.52–2.25) | 1.46 (1.27–1.68) |
| 5 - Least deprived (ref) | 1.00 | 1.00 |

Rows for deprivation groups 1 to 4 are adjusted odds ratios comparing that group with deprivation group 5 separately among children in care and children not in care. Estimates are derived from model (3), which adjusted for age at review, sex, and ethnicity and included a care status by deprivation group interaction. aOR = adjusted odds ratio.

### Appendix C.4 Sensitivity Analysis - Care-Status Differences in Psychosocial Health by Deprivation

Figure A6: Sensitivity analysis: Odds ratios comparing children in care with children not in care across psychosocial outcomes

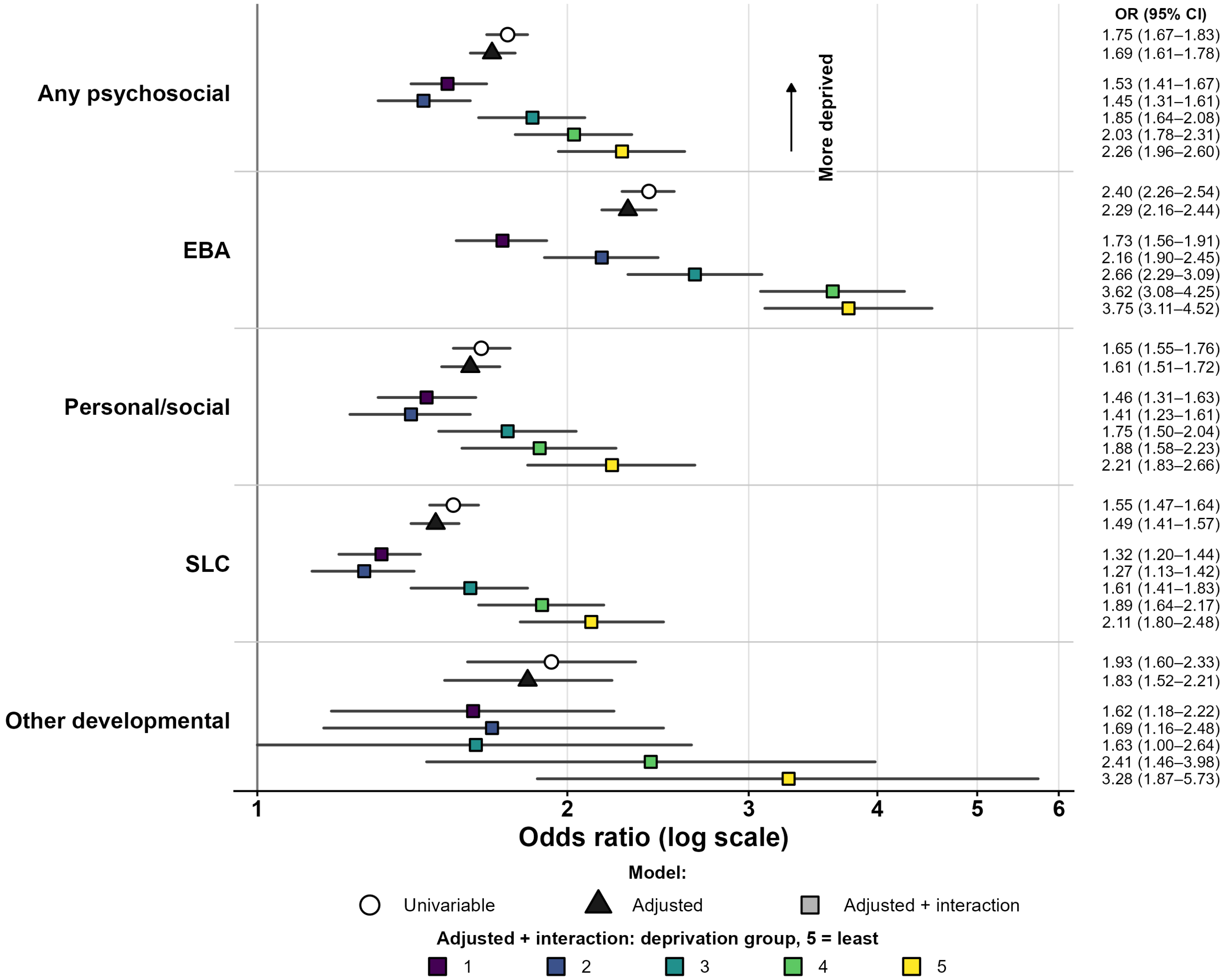


The "all potential cases" definition used in sensitivity analyses includes indicators rated *definite*, *probable*, or *possible*. Points show odds ratios and lines show 95% confidence intervals. The figure shows estimates from (1) unadjusted models, (2) adjusted models without interaction, and (3) adjusted models including a care status by deprivation group interaction. For model (3), estimates are odds ratios for children in care compared with children not in care within each deprivation group. Deprivation group 1 is the most deprived fifth of areas and group 5 is the least deprived fifth. EBA = emotional, behavioral, and/or attentional. SLC = speech, language, and/or communication.
