## Supplementary material for "Psychosocial Health Inequalities and Socioeconomic Deprivation Gradients Among Preschool Children in Care and Not in Care: An Administrative Health Data Study": RECORD guideline

### RECORD checklist

| Item | Reporting requirement | Location in manuscript / response |
|---|---|---|
| | **Title and abstract** | |
| 1a | State the study design using a commonly used term. | Abstract - Design: population-based cross-sectional study. |
| 1b | Provide an informative and balanced summary of what was done and found. | Abstract. |
| RECORD 1.1 | State the type of routinely collected data used; where possible, name the databases. | Title mentions administrative health data study. Abstract and Methods - Data sources identify the 27–30 Month Health Review and Scottish Birth Record data. |
| RECORD 1.2 | State the geographic region and timeframe, where applicable. | Abstract - Design and Abstract - Setting: Scotland; April 2013 to March 2023. |
| RECORD 1.3 | Clearly state any database linkage, where applicable. | Abstract - Design; Methods - Data sources. |
| | **Introduction** | |
| 2 | Explain the scientific background and rationale. | Introduction. |
| 3 | State specific objectives, including prespecified hypotheses where applicable. | Introduction - Study aims and research questions. |
| | **Methods** | |
| 4 | Present key elements of study design early in the paper. | Abstract - Design; Methods - Participants and Methods - Data sources. |
| 5 | Describe the setting, locations, and relevant dates. | Abstract - Design and Setting; Methods - Participants and Methods - Data sources. |
| 6a | Give eligibility criteria, sources and methods of participant selection, and follow-up methods where applicable. | Methods - Participants and Methods - Data sources; Results - Cohort characteristics. This is a cross-sectional study, so follow-up is not applicable. |
| 6b | For matched studies, give matching criteria and numbers exposed and unexposed. | Not applicable: the study was not matched. |
| RECORD 6.1 | Provide detailed information on the methods used to select the study population, including codes or algorithms where applicable. | Methods - Participants and Methods - Measures; Appendix A, especially care-status Read v2 codes and outcome-classification rules. |
| RECORD 6.2 | Cite validation studies of codes or algorithms used to select the population, where available. | No validation studies were available. The care-status and outcome definitions are described in Methods - Measures and Appendix A; all outcome classifications were checked by a medical doctor. |
| RECORD 6.3 | Describe the linkage of databases, including linkage methods and linkage quality. | Methods - Data sources: eDRIS performed all linkage and supplied access to a pseudonymized linked dataset. Linkage methods and linkage-quality metrics were not provided to the study team. |
| 7 | Clearly define outcomes, exposures, predictors, confounders, effect modifiers, and diagnostic criteria where applicable. | Methods - Measures and Methods - Statistical analysis and reporting; Appendix A. |
| RECORD 7.1 | Provide a complete list of codes and algorithms used to classify exposures, outcomes, confounders, and effect modifiers, or explain why this is not possible. | Appendix A provides outcome and care-status classification rules and Read v2 code lists. Demographic and deprivation variables are described in Methods - Data sources and Methods - Measures. |

| Item | Reporting requirement | Location in manuscript / response |
|---|---|---|
| 8 | For each variable, give sources of data and details of measurement; describe comparability across groups where relevant. | Methods - Data Sources and Methods - Measures; Appendix A. |
| 9 | Describe efforts to address potential sources of bias. | Methods - Participants, Methods - Data sources, Methods - Measures and Methods - Statistical analysis and reporting; Discussion - Limitations. |
| 10 | Explain how study size was arrived at. | Methods - Participants: all eligible records in the available extract were included; no sample-size calculation was performed. |
| 11 | Explain handling of quantitative variables and any groupings. | Methods - Measures and Methods - Statistical analysis and reporting. Age was continuous; SIMD was grouped into ordered quintiles. |
| 12a | Describe all statistical methods, including methods used to control confounding. | Methods - Statistical analysis and reporting. |
| 12b | Describe methods used to examine subgroups and interactions. | Methods - Statistical analysis and reporting: care-status by deprivation interaction models; Results - RQ2 and RQ3. |
| 12c | Explain how missing data were addressed. | Methods - Measures and Methods - Statistical analysis and reporting; Results - Cohort characteristics. Complete-case analysis was used for adjusted models. |
| 12d | For cohort studies, explain loss to follow-up; for case-control studies, matching; and for cross-sectional studies, the analytical approach. | Methods - Statistical analysis and reporting. Cross-sectional analytical approach described; loss to follow-up and matching not applicable. |
| 12e | Describe sensitivity analyses. | Methods - Statistical Analysis and Reporting; Results - RQ1 and RQ2; Appendix B; Appendix C.4. |
| RECORD 12.1 | Describe the extent of access to the database population used to create the study population. | Methods - Data Sources: eDRIS performed the linkage and provided the study team access only to a pseudonymized linked dataset. Acknowledgments: the secure analytical platform was within the National Safe Haven. |
| RECORD 12.2 | Provide information on data-cleaning methods. | Methods - Participants, Methods - Measures, and Methods - Statistical analysis and reporting. These sections describe eligibility restrictions, recoding of care status and ethnicity, deprivation-group assignment, and outcome derivation. |
| RECORD 12.3 | State whether the study included data linkage and describe linkage methods and linkage-quality evaluation. | Methods - Data sources. Linkage was performed by eDRIS; linkage methods and quality metrics were not provided to the study team. |
| | **Results** | |
| 13a | Report numbers of individuals at each stage of the study. | Results - Cohort characteristics: initial, age-eligible, known care-status, complete-covariate, and analytical cohorts. |
| 13b | Give reasons for non-participation at each stage. | Not applicable as a participation/recruitment study; exclusions due to eligibility and missing covariates are reported in Results - Cohort characteristics. |
| 13c | Consider use of a flow diagram. | Results - Cohort characteristics reports the participant flow numerically; a flow diagram was not included. |

| Item | Reporting requirement | Location in manuscript / response |
|---|---|---|
| RECORD 13.1 | Describe the selection of individuals, including filtering based on data quality, availability, and linkage. | Results - Cohort characteristics; Methods - Participants and Methods - Statistical analysis and reporting. |
| 14a | Give characteristics of study participants and information on exposures and potential confounders. | Table 1 and Results - Cohort characteristics. |
| 14b | Indicate the number of participants with missing data for each variable of interest. | Table 1; Results - Cohort characteristics. |
| 14c | For cohort studies, summarise follow-up time. | Not applicable: cross-sectional study. |
| 15 | Report numbers of outcome events or summary measures over time. | Results - RQ1–RQ3; Tables 2–3; Appendix B. |
| 16a | Give unadjusted and, if applicable, confounder-adjusted estimates with precision. | Results - RQ1–RQ3; Tables 2 and 3; Appendix B; Appendix C.1–C.3. |
| 16b | Report category boundaries when continuous variables were categorized. | Methods - Measures: SIMD quintiles; tables and figure legends. |
| 16c | Consider translating relative-risk estimates into absolute risk for a meaningful time period. | Not applicable: cross-sectional prevalence estimates are reported directly alongside relative measures. |
| 17 | Report other analyses, including subgroup, interaction, and sensitivity analyses. | Results - RQ1–RQ3; Appendix B; Appendix C.1–C.4. |
| | **Discussion** | |
| 18 | Summarise key results with reference to study objectives. | Discussion first paragraph and Discussion - Conclusion. |
| 19 | Discuss limitations, including potential direction and magnitude of bias. | Discussion - Limitations. |
| RECORD 19.1 | Discuss implications of using data not created to answer the research question, including misclassification, unmeasured confounding, missing data, and changing eligibility over time where relevant. | Discussion - Limitations: secondary data and recording practices, area-level deprivation, unavailable confounders, care-status recording, outcome classification, and generalisability. |
| 20 | Give a cautious overall interpretation considering objectives, limitations, analyses, and other evidence. | Discussion. |
| 21 | Discuss generalisability. | Discussion - Limitations and Discussion - Conclusion. |
| | **Other information** | |
| 22 | Give the source of funding and the role of funders. | Declarations, Funding and Role of the funders statements in back matter. |
| RECORD 22.1 | Provide information on access to supplementary information, study protocol, raw data, and programming code. | The manuscript includes Appendices A–C, and the completed RECORD checklist is supplied separately. No protocol or study plan was created. Declarations - Data Availability describes the controlled-access route for sensitive individual-level data. Declarations - Code Availability states that analysis code is available from the corresponding author on reasonable request. |
